# Effectiveness of SARS-CoV-2 mRNA Vaccines for Preventing Covid-19 Hospitalizations in the United States

**DOI:** 10.1101/2021.07.08.21259776

**Authors:** Mark W. Tenforde, Manish M. Patel, Adit A. Ginde, David J. Douin, H. Keipp Talbot, Jonathan D. Casey, Nicholas M. Mohr, Anne Zepeski, Manjusha Gaglani, Tresa McNeal, Shekhar Ghamande, Nathan I. Shapiro, Kevin W. Gibbs, D. Clark Files, David N. Hager, Arber Shehu, Matthew E. Prekker, Heidi L. Erickson, Matthew C. Exline, Michelle N. Gong, Amira Mohamed, Daniel J. Henning, Jay S. Steingrub, Ithan D. Peltan, Samuel M. Brown, Emily T. Martin, Arnold S. Monto, Akram Khan, C. Terri Hough, Laurence Busse, Caitlin C. ten Lohuis, Abhijit Duggal, Jennifer G. Wilson, Alexandra June Gordon, Nida Qadir, Steven Y. Chang, Christopher Mallow, Hayley B. Gershengorn, Hilary M. Babcock, Jennie H. Kwon, Natasha Halasa, James D. Chappell, Adam S. Lauring, Carlos G. Grijalva, Todd W. Rice, Ian D. Jones, William B. Stubblefield, Adrienne Baughman, Kelsey N. Womack, Christopher J. Lindsell, Kimberly W. Hart, Yuwei Zhu, Samantha M. Olson, Meagan Stephenson, Stephanie J. Schrag, Miwako Kobayashi, Jennifer R. Verani, Wesley H. Self, For the Influenza and Other Viruses in the Acutely Ill (IVY) Network

## Abstract

**Background:** As SARS-CoV-2 vaccination coverage increases in the United States (US), there is a need to understand the real-world effectiveness against severe Covid-19 and among people at increased risk for poor outcomes.

**Methods:** In a multicenter case-control analysis of US adults hospitalized March 11 - May 5, 2021, we evaluated vaccine effectiveness to prevent Covid-19 hospitalizations by comparing odds of prior vaccination with an mRNA vaccine (Pfizer-BioNTech or Moderna) between cases hospitalized with Covid-19 and hospital-based controls who tested negative for SARS-CoV-2.

**Results:** Among 1210 participants, median age was 58 years, 22.8% were Black, 13.8% were Hispanic, and 20.6% had immunosuppression. SARS-CoV-2 lineage B.1.1.7 was most common variant (59.7% of sequenced viruses). Full vaccination (receipt of two vaccine doses ≥14 days before illness onset) had been received by 45/590 (7.6%) cases and 215/620 (34.7%) controls. Overall vaccine effectiveness was 86.9% (95% CI: 80.4 to 91.2%). Vaccine effectiveness was similar for Pfizer-BioNTech and Moderna vaccines, and highest in adults aged 18-49 years (97.3%; 95% CI: 78.9 to 99.7%). Among 45 patients with vaccine-breakthrough Covid hospitalizations, 44 (97.8%) were ≥50 years old and 20 (44.4%) had immunosuppression. Vaccine effectiveness was lower among patients with immunosuppression (59.2%; 95% CI: 11.9 to 81.1%) than without immunosuppression (91.3%; 95% CI: 85.5 to 94.7%).

**Conclusion:** During March–May 2021, SARS-CoV-2 mRNA vaccines were highly effective for preventing Covid-19 hospitalizations among US adults. SARS-CoV-2 vaccination was beneficial for patients with immunosuppression, but effectiveness was lower in the immunosuppressed population.

## INTRODUCTION

Over 2.2 million hospitalizations and 580,000 deaths related to coronavirus disease 2019 (Covid-19) occurred in the United States (US) through May 2021.[1] In December 2020, the Food and Drug Administration granted Emergency Use Authorization (EUA) for two messenger RNA (mRNA) vaccines (from Pfizer-BioNTech and Moderna) against severe acute respiratory syndrome coronavirus 2 (SARS-CoV-2),[2] the virus that causes Covid-19. Widespread public health initiatives resulted in over 60% of the US adult population receiving at least one dose of a SARS-CoV-2 vaccine by the end of May 2021.[1] The mRNA vaccines have been the predominate SARS-CoV-2 vaccine products used in the US, with approximately 95% of vaccinated people having received either a Pfizer-BioNTech or Moderna vaccine.[1]

Phase 3 clinical trials of mRNA vaccines found a 94–95% reduction in Covid-19 illness and near 100% protection against severe Covid-19.[3, 4] However, these clinical trials had few cases of hospitalized Covid-19, and limited power to assess efficacy among persons with underlying illnesses who are at high risk for severe Covid-19. As vaccine coverage increases, observational vaccine effectiveness evaluations are important to understand how well the vaccines protect against Covid-19 in real-world settings across diverse populations, including immunocompromised hosts. The Centers for Disease Control and Prevention (CDC) collaborates with the Influenza and Other Viruses in the Acutely Ill (IVY) Network to monitor the effectiveness of SARS-CoV-2 vaccines for the prevention of Covid-19 hospitalizations among US adults. In this analysis, we evaluated the effectiveness of SARS-CoV-2 mRNA vaccines for preventing Covid-19 hospitalizations by vaccine product, by age group, and by underlying medical conditions, including immunosuppression.[5]

## METHODS

### Design

The protocol and statistical analysis plan for this project are available in the supplementary materials. We conducted a prospective observational case-control evaluation of vaccine effectiveness by comparing the odds of antecedent SARS-CoV-2 vaccination in hospitalized case-patients with Covid-19 versus control-patients without Covid-19. We included two control groups: 1) “test-negative” controls presented with signs or symptoms of an acute respiratory illness but tested negative for SARS-CoV-2; and 2) “syndrome-negative” controls were selected from hospitalized adults without signs or symptoms of an acute respiratory illness and tested negative for SARS-CoV-2. Test-negative controls are commonly used in hospital-based vaccine effectiveness evaluations;[6-9] in this test-negative design, utilizing a comparison group with the same clinical syndrome and similar level of acuity as cases reduces bias due to differential healthcare seeking behavior. Because of the potential for misclassification of true cases as test-negative controls due to false-negative test results using the test-negative design, particularly for those presenting late in the course of illness,[10] we included the second control group of hospitalized patients without an acute respiratory illness. Consistent with methodologies for vaccine effectiveness evaluations recommended by the World Health Organization,[11] SARS-CoV-2 vaccine coverage in both control groups served as a proxy for background vaccination rates in the source population for COVID-19 cases.

### Setting

This surveillance activity was conducted by the IVY Network, a CDC-funded collaborative of hospital-based investigators across the US,[6, 12] and included patients hospitalized from March 11 through May 5, 2021, at 18 academic medical centers in 16 states. Each participating institution, including CDC, reviewed and conducted this activity consistent with applicable federal law and CDC policy (45 C.F.R. part 46.102(l)(2), 21 C.F.R. part 56; 42 U.S.C. §241(d); 5 U.S.C. §552a; 44 U.S.C. §3501 et seq.). The program was conducted as a public health surveillance activity without written informed consent.

### Participants

Sites screened hospitalized adults ≥18 years old for potential eligibility through daily review of hospital admission logs and electronic medical records. Detailed eligibility criteria are shown in Supplementary Appendix B. In brief, Covid-19 cases included patients with a clinical syndrome consistent with acute Covid-19 and a positive test for SARS-CoV-2 within 10 days following symptom onset.[13-15] Test-negative controls had a clinical syndrome consistent with acute Covid-19 but tested negative for SARS-CoV-2. Syndrome-negative controls did not have a clinical syndrome consistent with Covid-19 and tested negative for SARS-CoV-2. Individual matching between cases and controls was not performed. Sites targeted a case: control ratio of approximately 1:1 for each control group and did not seek information on vaccination status until after patients were included.

### Data Collection

Participants (or their proxies when participants could not answer medical questions) were interviewed by trained personnel to collect data on demographics, medical conditions, SARS-CoV-2 vaccination, and other patient characteristics. Additional information on underlying medical conditions and SARS-CoV-2 clinical testing was obtained through standardized medical record review.

### Laboratory Analysis

Upper respiratory specimens (nasal swabs or saliva) were collected, frozen, and shipped to a central laboratory at Vanderbilt University Medical Center (Nashville, Tennessee). Specimens underwent reverse transcription polymerase chain reaction (RT-PCR) testing for the SARS-CoV-2 nucleocapsid gene using standardized methods and interpretive criteria.[16] Specimens positive for SARS-CoV-2 with a cycle threshold <32 for at least one of the two RT-PCR targets were shipped to the University of Michigan (Ann Arbor, Michigan) for viral whole genome sequencing using the ARTIC Network version 3 protocol on an Oxford Nanopore Technologies instrument (Supplementary Appendix B).[17] SARS-CoV-2 lineages were assigned with >80% coverage using Pangolin genomes.[18]

### Classification of Case-Control Status

Final classification of case-control status was determined with consideration of both clinical SARS-CoV-2 testing completed at local hospital laboratories and RT-PCR testing completed at the central laboratory. Cases tested positive for SARS-CoV-2 by a clinical test or central laboratory RT-PCR test. Cases with SARS-CoV-2 detected by RT-PCR with a cycle threshold >32 were included in the analysis, but viral sequencing information was not available for these cases. Test-negative controls tested negative for SARS-CoV-2 by all clinical and central testing. Patients initially classified as a test-negative control who subsequently tested positive for SARS-CoV-2 by central RT-PCR testing were reclassified as cases. Syndrome-negative controls tested negative for SARS-CoV-2 by all clinical and central testing. Patients included as syndrome-negative controls who subsequently tested positive for SARS-CoV-2 in the central laboratory were excluded from the analysis.

### Classification of Vaccination Status

Details of SARS-CoV-2 vaccination, including dates and location of vaccination, vaccine product, and lot number, were ascertained through a systematic process including patient or proxy interview and source verification. Sources of documentation included vaccination card, hospital records, state vaccine registries which were searched at the time of interview and again approximately 28 days later, and vaccine records requested from clinics and pharmacies. Vaccine doses were classified as administered if source documentation was identified or if the patient/proxy reported a vaccine dose with a plausible date and location of vaccination.

The SARS-CoV-2 mRNA vaccines are administered as a two-dose series; participants were considered fully vaccinated 14 days after receipt of the second vaccine dose.[19] Vaccination status was classified based on the number of mRNA vaccine doses received before a reference date, which was the date of symptom onset for cases and test-negative controls and date of hospital admission for syndrome-negative controls. Participants were classified as: unvaccinated if they had received no vaccine doses prior to the reference date; partially vaccinated if they received one dose ≥14 days before the reference date; and fully vaccinated if they received both doses ≥14 days before the reference date. As protective immunity from SARS-CoV-2 vaccines is not expected immediately after the first dose,[12] patients who received a first dose <14 days before the reference date were excluded from the analysis. Patients who received a SARS-CoV-2 vaccine that had not been authorized in the US were excluded. Due to recent introduction of the Janssen (Johnson & Johnson) SARS-CoV-2 vaccine following its EUA in February 2021,[2] patients who received this vaccine were also excluded.

### Statistical Analysis

Vaccine effectiveness and 95% confidence intervals (95% CI) were determined by comparing the odds of prior SARS-CoV-2 vaccination in case-patients and control-patients, calculated as: vaccine effectiveness = (1 – odds ratio) × 100%.[20]

Primary vaccine effectiveness estimates were calculated in adults of all ages for full vaccination versus unvaccinated and for partial vaccination versus unvaccinated. Unadjusted vaccine effectiveness was calculated with simple logistic regression and then a model building approach was applied to estimate adjusted vaccine effectiveness accounting for potential confounders. Prespecified covariates in base multivariable logistic regression models included calendar time in biweekly intervals, US Department of Health and Human Services region, age, sex, and self-reported race and Hispanic ethnicity. We repeated the regression by adding health status indicators and SARS-CoV-2 exposure variables potentially associated with the likelihood of vaccination and risk of Covid-19 hospitalization (detailed in Supplementary Appendix B). No variable generated an absolute change in the odds ratio of vaccination of more than 5% in either direction, which was a prespecified cutoff, so no additional variables were added to the base model. Potential effect modification of prior SARS-CoV-2 infection (at least 14 days prior to the current illness) was assessed using a likelihood ratio test (with a P-value <0.15 suggestive of effect modification).[21]

Separate assessments were initially performed using the test-negative control and the syndrome-negative control group to assess comparability of estimates. Effectiveness estimates were very similar using the test-negative and syndrome-negative control groups. Therefore, control groups were combined to improve precision.

Vaccine effectiveness estimates were stratified by age group (18–49, 50–64, or ≥65 years), SARS-CoV-2 vaccine product (Pfizer-BioNTech or Moderna), SARS-CoV-2 variant, and underlying medical conditions with a prevalence ≥20% in the population, including immunocompromising conditions[6], diabetes mellitus, chronic lung disease, chronic cardiovascular disease, and obesity (definitions provided in Supplementary Appendix B). Sensitivity analyses are described in Supplementary Appendix B. Stata Version 16 (College Station, TX) and SAS 9.4 (Cary, NC) were used for statistical analysis.

## RESULTS

### Participants

We included 1210 patients (590 cases, 334 test-negative controls, and 286 syndrome-negative controls) (Figure S1) enrolled from 18 clinical sites (Figure 1) over the course of 51 days (Figure S2). Overall, median age was 58 years, 276 (22.8%) were non-Hispanic Black, 167 (13.8%) were Hispanic, and 248 (20.6%) had an immunocompromising condition (Table 1).

**Table 1.**
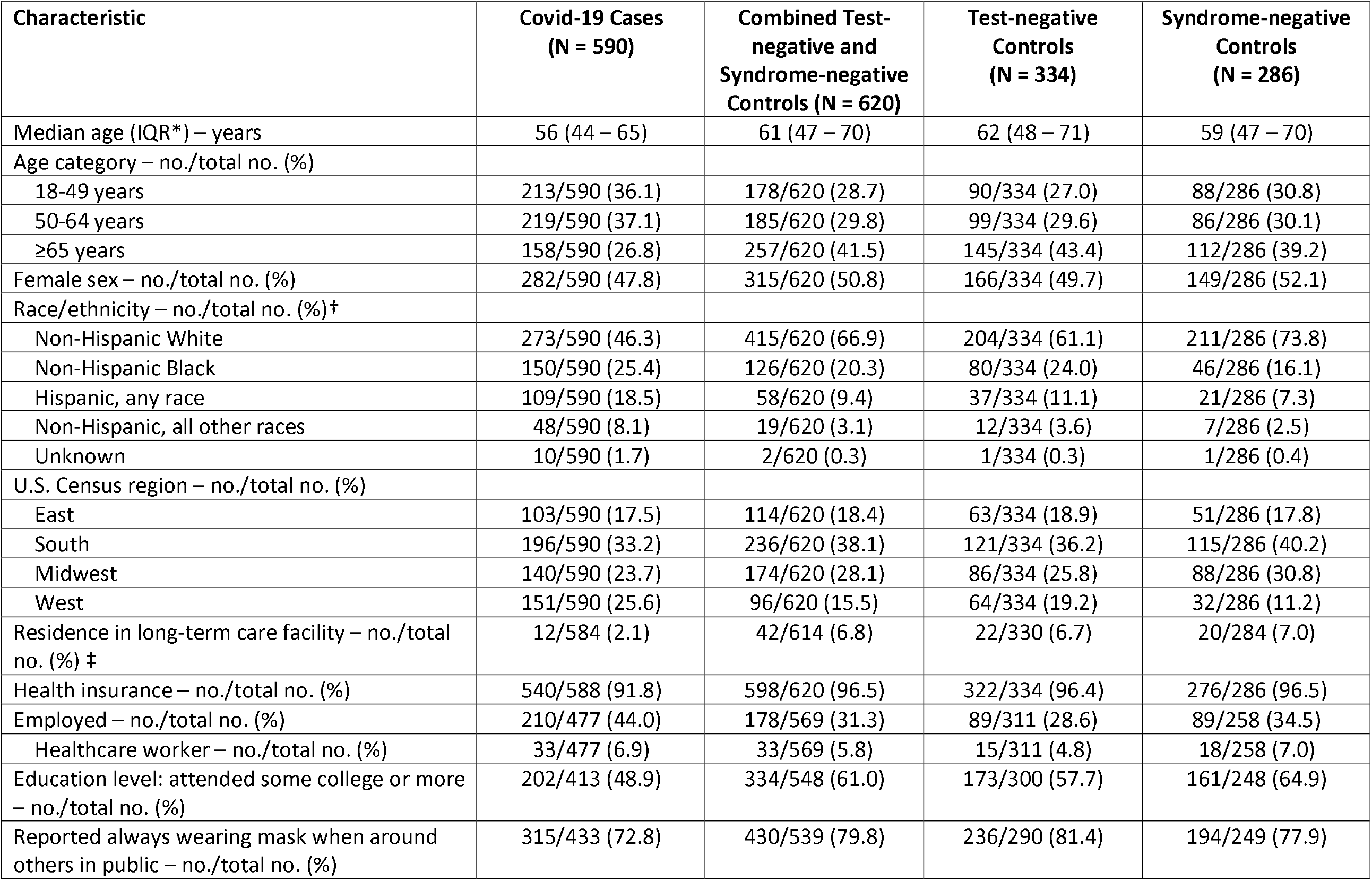
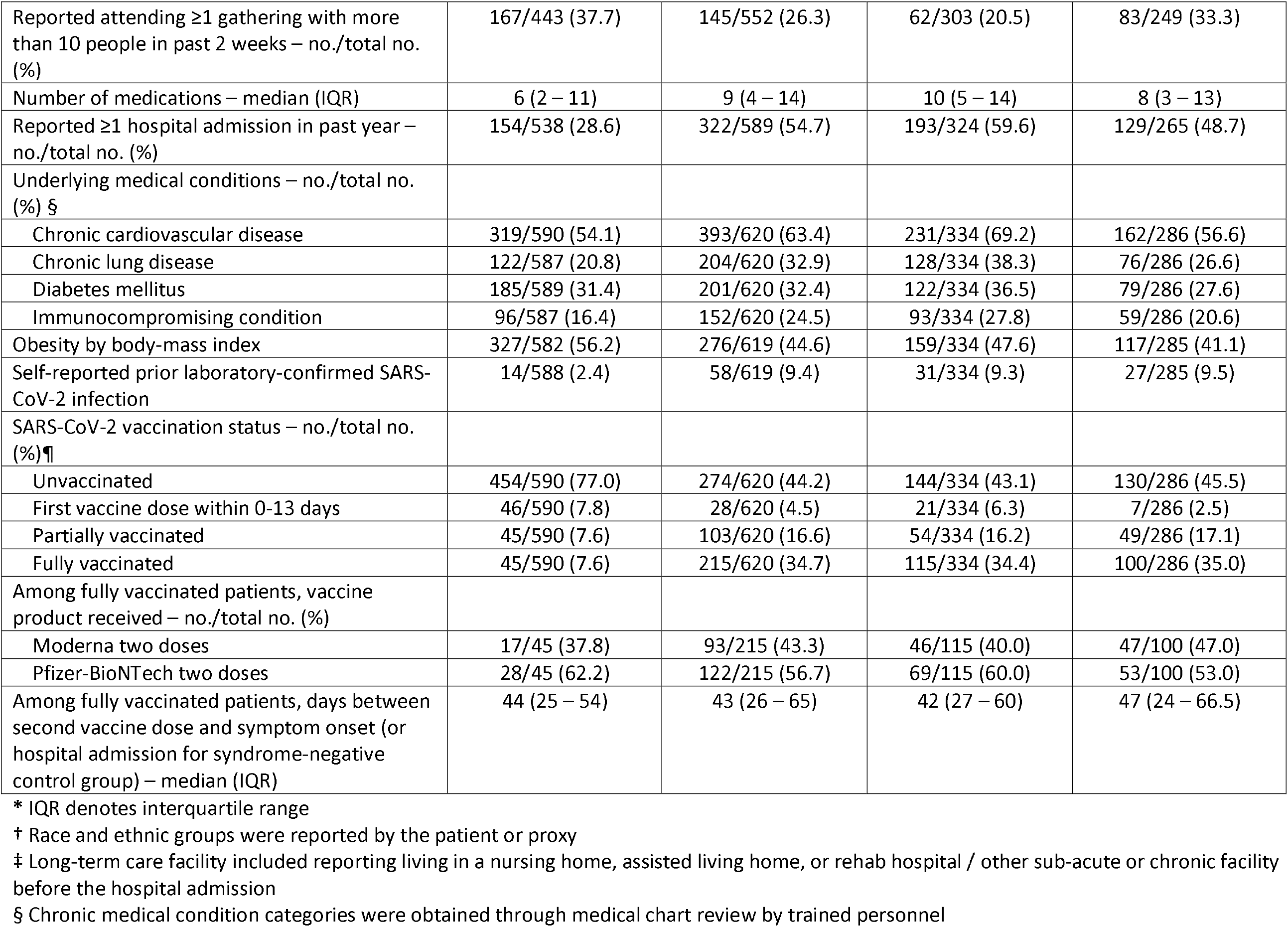
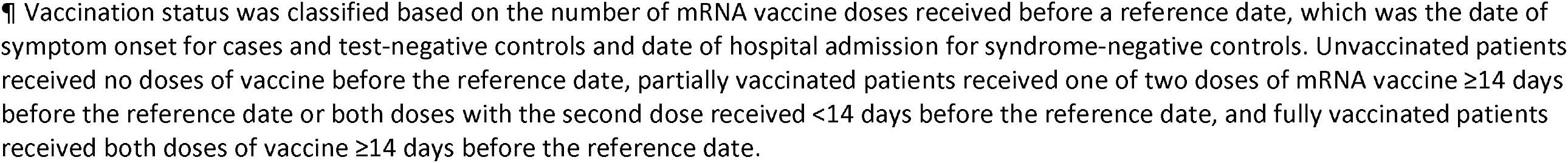
Characteristics of hospitalized Covid-19 case and control patients — IVY Network, United States, March–May 2021.

**Figure 1.**
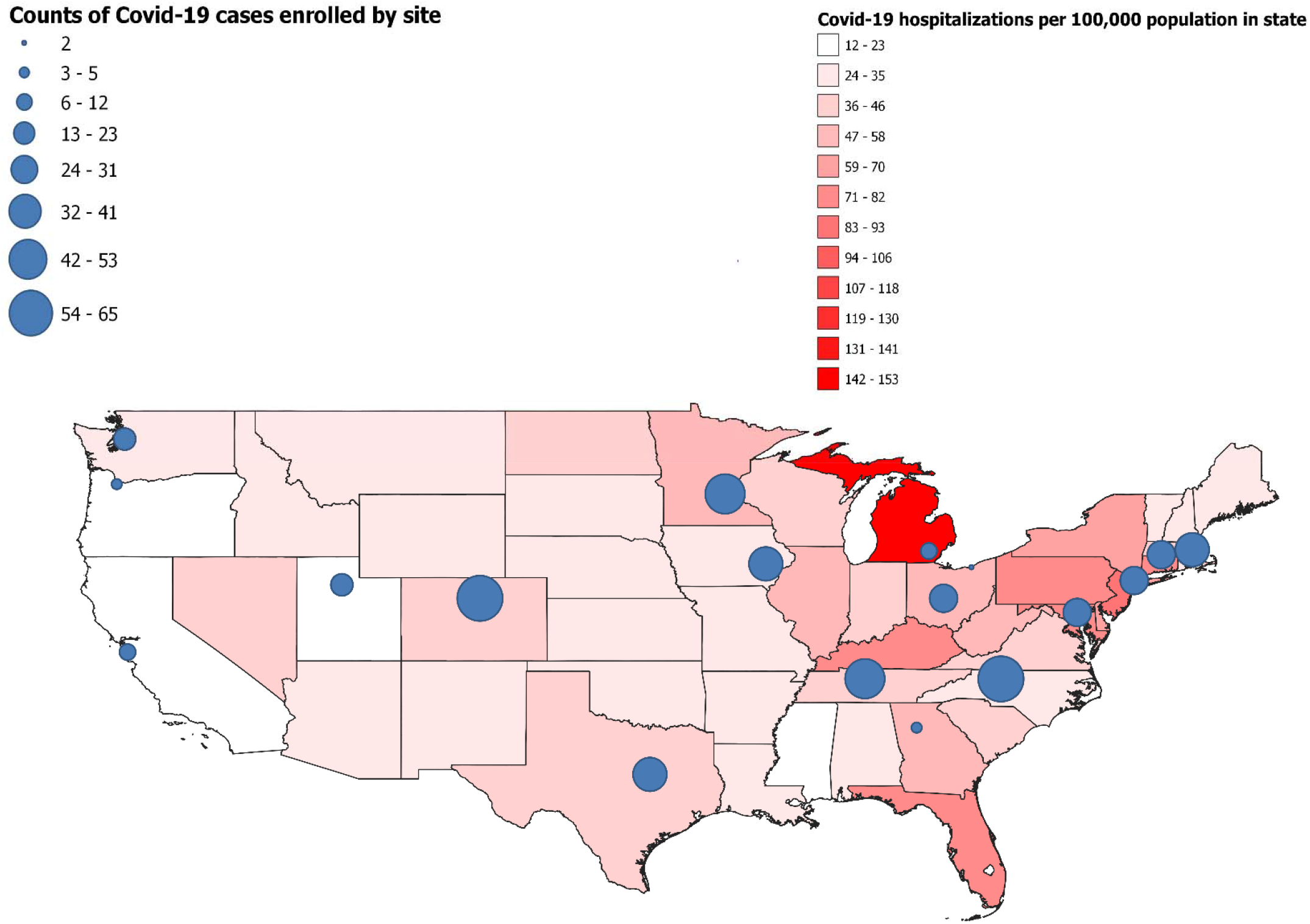
Map of continental United States with incidence of Covid-19 hospitalizations by state in April 2021 indicated by color (red). Participating sites are shown on the map with circles; the size of each circle represents the number of Covid-19 cases included from each site in this analysis -- IVY Network, United States, March–May 2021.* ^*^ Sources of state Covid-19 hospitalization data and population census data were HealthData.gov and United States Census Bureau. [31, 32]

After excluding patients with a first vaccine dose 0-13 days before the reference date, full SARS-CoV-2 vaccination had been received by 45 (8.3%) cases, 115 (36.7%) test-negative controls, and 100 (35.6%) syndrome-negative controls (Figure 2); 454 (83.5%) cases were unvaccinated. Among fully vaccinated patients, median time between the last vaccine dose and onset of Covid-like symptoms was 44 days (interquartile range [IQR] 25 to 54 days) for 45 cases and 42 days (IQR 27 to 60 days) for 115 test-negative controls. Among fully vaccinated patients, 251 (96.5%) had source verification of vaccine doses. SARS-CoV-2 whole genome sequencing was completed for 231 cases, with variant of concern B.1.1.7 the most commonly identified lineage (138/231, 59.7%) (Table 2).

**Table 2.**
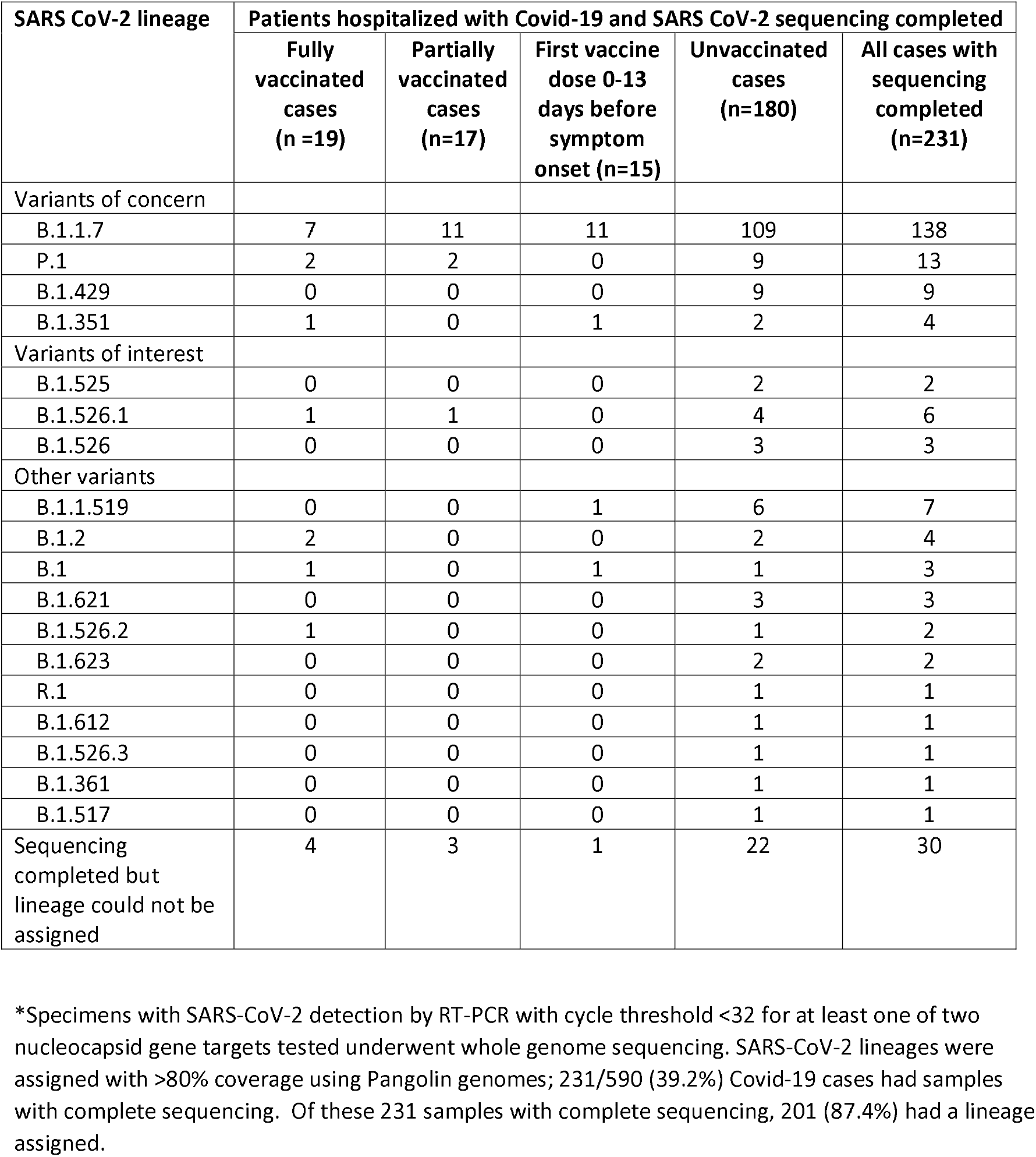
SARS-CoV-2 lineages identified by whole genome sequencing of upper respiratory specimens collected from Covid-19 cases — IVY Network, United States, March–May 2021.*

| SARS CoV-2 lineage | Patients hospitalized with Covid-19 and SARS CoV-2 sequencing completed |  |  |  |  |
| --- | --- | --- | --- | --- | --- |
|  | Fully vaccinated cases (n=19) | Partially vaccinated cases (n=17) | First vaccine dose 0-13 days before symptom onset (n=15) | Unvaccinated cases (n=180) | All cases with sequencing completed (n=231) |
| Variants of concern |  |  |  |  |  |
| B.1.1.7 | 7 | 11 | 11 | 109 | 138 |
| P.1 | 2 | 2 | 0 | 9 | 13 |
| B.1.429 | 0 | 0 | 0 | 9 | 9 |
| B.1.351 | 1 | 0 | 1 | 2 | 4 |
| Variants of interest |  |  |  |  |  |
| B.1.525 | 0 | 0 | 0 | 2 | 2 |
| B.1.526.1 | 1 | 1 | 0 | 4 | 6 |
| B.1.526 | 0 | 0 | 0 | 3 | 3 |
| Other variants |  |  |  |  |  |
| B.1.1.519 | 0 | 0 | 1 | 6 | 7 |
| B.1.2 | 2 | 0 | 0 | 2 | 4 |
| B.1 | 1 | 0 | 1 | 1 | 3 |
| B.1.621 | 0 | 0 | 0 | 3 | 3 |
| B.1.526.2 | 1 | 0 | 0 | 1 | 2 |
| B.1.623 | 0 | 0 | 0 | 2 | 2 |
| R.1 | 0 | 0 | 0 | 1 | 1 |
| B.1.612 | 0 | 0 | 0 | 1 | 1 |
| B.1.526.3 | 0 | 0 | 0 | 1 | 1 |
| B.1.361 | 0 | 0 | 0 | 1 | 1 |
| B.1.517 | 0 | 0 | 0 | 1 | 1 |
| Sequencing completed but lineage could not be assigned | 4 | 3 | 1 | 22 | 30 |

\*Specimens with SARS-CoV-2 detection by RT-PCR with cycle threshold <32 for at least one of two nucleocapsid gene targets tested underwent whole genome sequencing. SARS-CoV-2 lineages were assigned with >80% coverage using Pangolin genomes; 231/590 (39.2%) Covid-19 cases had samples with complete sequencing. Of these 231 samples with complete sequencing, 201 (87.4%) had a lineage assigned.

**Figure 2.**
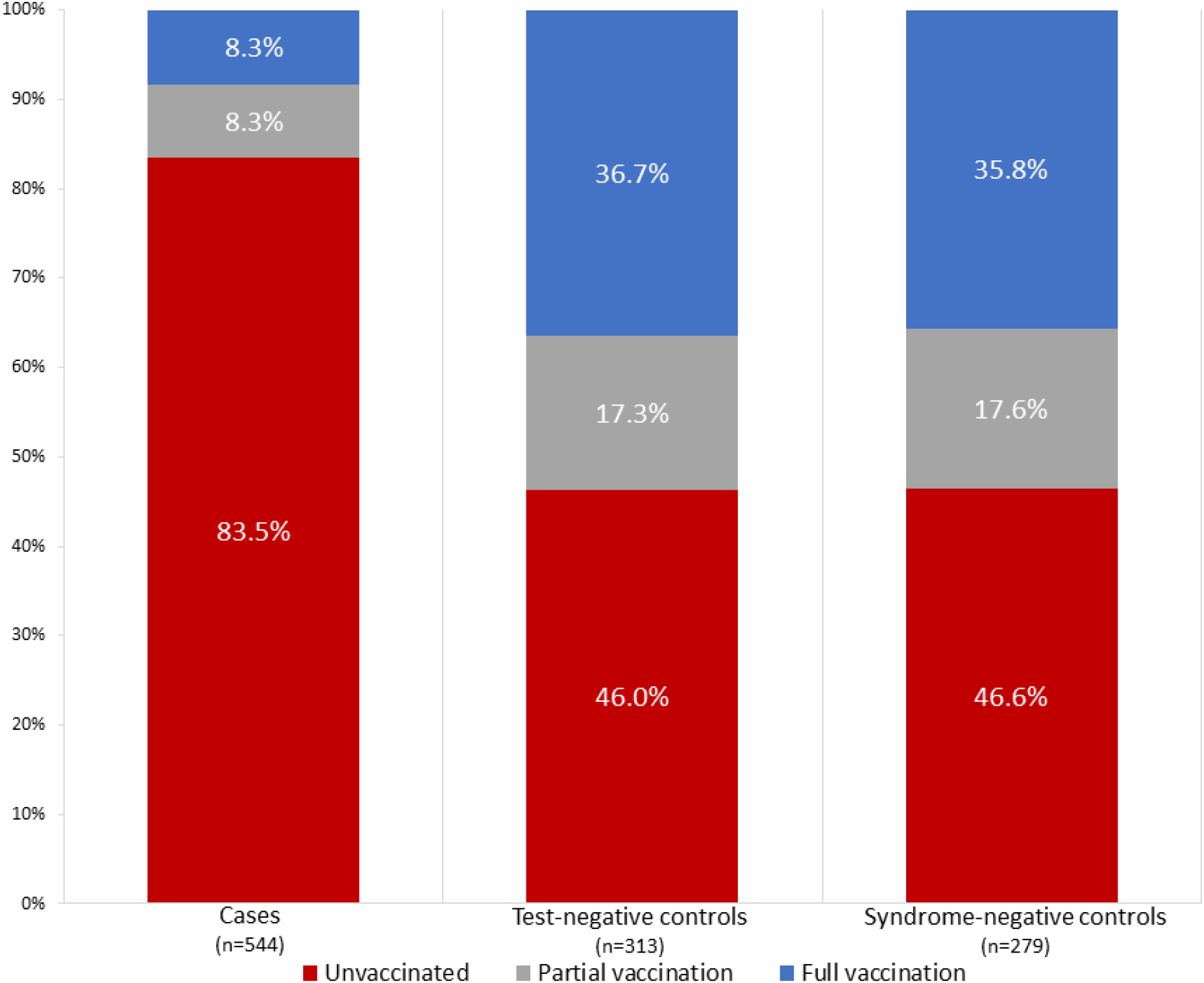
Vaccination status of case patients (N=544), test-negative controls (N=313), and syndrome-negative controls (N=279) — IVY Network, United States, March–May 2021.^*^ ^*^ Vaccination status was classified based on the number of mRNA vaccine doses received before a reference date, which was the date of symptom onset (for cases and test-negative controls) or date of hospital admission (for syndrome-negative controls). Unvaccinated patients received no doses of mRNA vaccine before the reference date, partially vaccinated patients received one of two doses of mRNA vaccine ≥14 days before the reference date or both doses with the second dose received <14 days before the reference date, and fully vaccinated patients received both doses of vaccine ≥14 days before the reference date. Patients who received the first dose of a SARS-CoV-2 vaccine 0-13 days before the reference date (46 cases; 21 test-negative controls; 7 syndrome-negative controls) were excluded from this analysis.

### Vaccine effectiveness

Vaccine effectiveness results for full vaccination were very similar using the test-negative control group (86.4%, 95% CI: 78.7–91.3%) and syndrome-negative control group (86.8%, 95% CI: 78.7–91.8%) (Figure S3 and Figure S4). After combining control groups, SARS-CoV-2 vaccine effectiveness for full vaccination to prevent Covid-19 hospitalizations was 86.9% (95% CI: 80.4–91.2%) (Figure 3). Vaccine effectiveness for full vaccination was similar for Pfizer-BioNTech (84.3%, 95% CI: 74.6–90.3%) and Moderna (90.0%, 95% CI: 82.0–94.4) vaccines. Point estimates were higher for people aged 18–49 years (97.3%, 95% CI: 78.9–99.7%) than aged 50–64 years (74.7%, 95% CI: 47.2–87.9%) and aged ≥65 years (87.2%, 95% CI: 77.6–92.7%). Among adults aged ≥65 years, vaccine effectiveness was similar in those 65-74 years (87.9%, 95% CI: 74.0–94.4%) and ≥75 years (90.5%, 95% CI: 73.2–96.7%). Vaccine effectiveness against SARS-CoV-2 B.1.1.7 lineage was 92.8% (95% CI: 83.0–96.9%).

**Figure 3.**
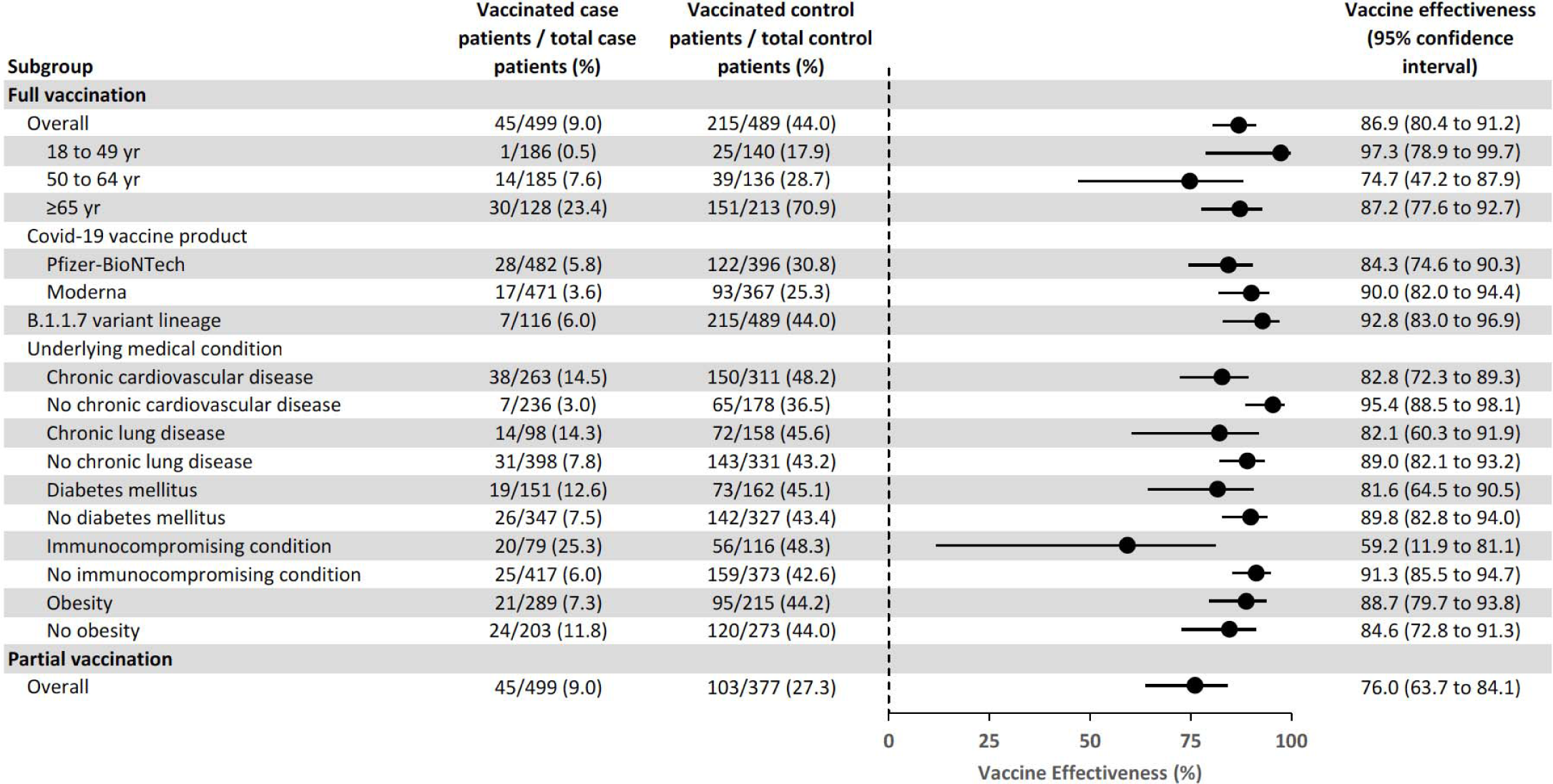
Vaccine effectiveness of SARS-CoV-2 mRNA vaccines for the prevention of Covid-19 hospitalizations overall and by subgroups — IVY Network, United States, March–May 2021.^*^ ^*^ The analysis included case patients with Covid-19-like illness who tested positive for SARS-CoV-2 infection and control patients combined from two groups, including a (1) test-negative control group with Covid-19-like illness and negative SARS-CoV-2 testing and (2) a syndrome-negative control group without Covid-19-like illness and negative for SARS-CoV-2. Vaccine effectiveness models were adjusted for calendar time in biweekly intervals, US Department of Health and Human Services region, age in years, sex, race and ethnicity. Vaccination status was classified based on the number of mRNA vaccine doses received before a reference date, which was defined as the date of symptom onset for cases and test-negative controls and date of hospital admission for syndrome-negative controls. Unvaccinated patients received no doses of vaccine before the reference date, partially vaccinated patients received one of two doses of vaccine ≥14 days before the reference date or both doses with the second dose received <14 days before the reference date, and fully vaccinated patients received both doses of vaccine ≥14 days before the reference date.

Vaccine effectiveness was significantly reduced for patients with immunocompromising conditions (59.2%, 95% CI: 11.9–81.1%) compared to individuals without an immunocompromising condition (91.3%, 95% CI: 85.5-94.7%) (Figure 3). Restricted to immunocompromised patients with an active solid organ or hematologic malignancy or solid organ transplant, vaccine effectiveness was 51.2% (95% CI: -30.7–81.8%). Vaccine effectiveness point estimates were lower for patients with underlying cardiovascular disease (82.8%, 95% CI: 72.3–89.3%), chronic lung disease (82.1%, 95% CI: 60.3–91.9%), and diabetes mellitus (81.6%, 95% CI: 64.5–90.5%) compared to patients without these underlying conditions but 95% confidence limits overlapped. Partial vaccination had a vaccine effectiveness of 76.0% (95% CI: 63.7–84.1%). Evidence of effect modification by prior laboratory-confirmed SARS-CoV-2 infection was not observed (likelihood ratio test p-value = 0.55). Sensitivity analyses produced results similar to the primary analysis (Table S2).

### Breakthrough vaccine Covid-19 hospitalizations

Forty-five Covid-19 case patients met our definition of being fully vaccinated before symptom onset. Among these, median age was 68 years (IQR 62–77 years), median time between the final vaccine dose and symptom onset was 44 days (IQR: 25 to 54 days), and 20 (44.4%) had an immunocompromising condition, including active solid organ or hematologic malignancy (n=9), and prior solid organ transplant (n=7). These breakthrough cases included 28 patients vaccinated with the Pfizer-BioNTech product and 17 patients vaccinated with the Moderna product.

## DISCUSSION

In this prospective observational surveillance program conducted at 18 geographically dispersed sites in the US during the early phase of the SARS-CoV-2 vaccination program, the two mRNA vaccines authorized for use in the US were approximately 87% effective for preventing Covid-19 hospitalizations, with similar effectiveness observed for the Pfizer BioNTech and Moderna products. This analysis adds to early real-world evaluations that demonstrated high vaccine effectiveness against Covid-19 in groups prioritized for early vaccination, such as healthcare workers.[22, 23] These results also add to a limited body of evidence that the SARS-CoV-2 mRNA vaccines are highly effective for preventing Covid-19 hospitalizations.

These results expand upon findings of high efficacy for SARS-CoV-2 mRNA vaccines reported from phase 3 clinical trials.[3, 4] The trials included patient populations healthier at baseline than those commonly hospitalized with Covid-19 and were not powered to evaluate the protective benefits of vaccination for preventing severe outcomes, such as hospitalization. The design of this surveillance analysis with concurrent inclusion of patients hospitalized with Covid-19 and two separate control groups enabled a robust evaluation of vaccine effectiveness for the prevention of severe Covid-19, including among patients with multiple and serious medical comorbidities. Vaccination coverage in the two control groups was very similar to one another and to vaccine uptake in the US during the surveillance period for this analysis,[1] adding confidence to our vaccine effectiveness results. The findings of high vaccine effectiveness in the US adult population and across subgroups defined by age, demographics, and comorbidities suggest that the mRNA vaccines are broadly effective for the prevention of severe Covid-19, including in populations at high risk of severe illness.

The protective benefits of any vaccination require an immune response to the vaccine. A history of solid organ transplant and other immunocompromising conditions have been associated with reduced cell-mediated and humoral immune responses to SARS-CoV-2 vaccines.[24, 25] Our results suggested substantial clinical benefit from vaccination in immunosuppressed people, with a vaccine effectiveness of about 60% for the prevention of Covid-19 hospitalizations in this population. However, vaccine effectiveness was significantly lower in patients with immunocompromising conditions compared to those without immunocompromising conditions. Among patients with vaccine breakthrough Covid-19 hospitalizations in this analysis, almost one-half had an immunocompromising condition, most commonly a history of solid organ transplantation or an actively treated malignancy. Immunosuppressive conditions affect millions of adults in the United States.[26] Future work is needed to understand vaccine effectiveness among people with specific immunocompromising conditions and the durability of protection in this population to inform the need for booster vaccines and/or non-vaccine preventative interventions, such as mask use and social distancing.

During implementation of the national SARS-CoV-2 vaccination program in Israel between December 2020 and February 2021, the Pfizer BioNTech vaccine product demonstrated 87% vaccine effectiveness for the prevention of Covid-19 hospitalizations with a mean follow-up of 15 days.[27] Results of our US analysis in a population with a higher burden of comorbidities demonstrated similar vaccine effectiveness for both the Pfizer BioNTech and Moderna mRNA vaccines with longer follow-up time (median 43 days and maximum 113 days). Evaluating the duration of protection from SARS-CoV-2 vaccines will require additional evaluation with longer follow-up time.

This analysis had certain limitations. While we included control groups that were likely to reduce bias from differential healthcare seeking behavior, there was potential for residual confounding. People who chose to be vaccinated may have been more likely to engage in other behaviors to reduce their risk for Covid-19, such as mask use and avoiding large crowds. Adjusting for self-reported variables on non-vaccine preventive measures did not substantively change vaccine effectiveness estimates. Race and Hispanic ethnicity differed between case and control groups; this likely represented underlying differences in the incidence of SARS-CoV-2 infection by race and ethnicity in the US and models were adjusted for race and ethnicity.[28] Although vaccine effectiveness was lower in adults with immunocompromising conditions, these conditions are likely associated with varying severity of immunosuppression and this analysis was not powered to look at vaccine effectiveness among subgroups with individual immunocompromising conditions. In an effort to capture all COVID-19 cases admitted to participating hospitals during a period of high community transmission, enrollment of cases and controls was not matched on a day-to-day basis; however, all cases and controls were enrolled within a 51-day period and vaccine effectiveness models were adjusted for calendar time. For certain exposures or behaviors such as mask use, there was a potential for recall bias or social desirability bias. However, inclusion of these variables in adjusted models did not substantively change vaccine effectiveness estimates and they were not included in the final models. Lastly, most sequenced viruses in this analysis were B.1.1.7 variants, which represented the majority of circulating viruses in the US during this time period;[1] vaccine effectiveness against other emerging variants will require additional study.

In conclusion, the SARS-CoV-2 mRNA vaccines were highly effective for preventing Covid-19 hospitalizations among adults in March through May 2021. Widespread vaccination can be expected to have a major beneficial impact on Covid-19 hospitalizations and associated outcomes, such as death and post-Covid complications.[29, 30] While SARS-CoV-2 mRNA vaccines appear to provide substantial benefit to immunocompromised people, effectiveness is lower in this population than in the immunocompetent population. It will be crucial to understand the benefit of additional preventive measures, such as vaccine boosters and continued masking, in patients at highest risk for vaccine breakthrough.

## Supporting information

Protocol and Statistical Analysis Plan

Strobe checklist

## Data Availability

Data from this project are not available outside of the investigator group.

## Funding

Primary funding for this study was provided by the US Centers for Disease Control and Prevention (75D30121F00002). The REDCap data tool was supported by a Clinical and Translational Science Award (UL1 TR002243) from the National Center for Advancing Translational Sciences.

## Potential Conflicts of Interest

Vaccine manufacturers had no role in the conduct, analysis, or dissemination of this work. The primary funder for this work was the US Centers for Disease Control and Prevention (CDC); CDC scientists participated in this work and are included as authors. The following potential conflicts of interest have been reported by the authors. Dr. Brown reports grants from CDC during the conduct of the study; personal fees from Hamilton, other from Faron, other from Sedana, grants from Janssen, grants from NIH, grants from DoD, other from Oxford University, other from Brigham Young University, outside the submitted work. Dr. Casey reports grants from National Institute of Health, outside the submitted work. Dr. Chang was a speaker for La Jolla Pharmaceuticals in 2018, and consulted for PureTech Health in 2020. Dr. Chappell reports grants from CDC, grants from NCATS/NIH during the conduct of the study. Dr. Exline reports other from Abbott Labs, outside the submitted work. Dr. Files reports personal fees as consultant from Cytovale, DSMB member from Medpace, outside the submitted work. Dr. Gaglani reports grants from CDC-Vanderbilt, during the conduct of the study; grants from CDC, grants from CDC, grants from CDC-Abt, grants from CDC-Westat, outside the submitted work. Dr. Gershengorn reports personal fees from Gilead Sciences, Inc, outside the submitted work. Dr. Ginde reports grants from CDC, during the conduct of the study; grants from AbbVie, grants from Faron Pharmaceuticals, outside the submitted work. Dr. Gong reports grants from CDC, during the conduct of the study; grants from NIH, grants from AHRQ, other from Regeneron, personal fees from Philips Healthcare, outside the submitted work. Dr. Grijalva reports other from Pfizer, other from Merck, other from Sanofi-Pasteur, grants from Campbell Alliance/Syneos Health, grants from Centers for Disease Control and Prevention, grants from National Institutes of Health, grants from Food and Drug Administration, grants from Agency for Health Care Research and Quality, grants from Sanofi, outside the submitted work. Dr. Hager reports other from CDC via subcontract with Vanderbilt during the conduct of the study; other from Incyte Corporation, other from Marcus Foundation, other from EMPACT Precision Medicine via VUMC, outside the submitted work. Dr. Halasa reports grants from CDC during the conduct of the study; grants and non-financial support from Sanofi, grants from Quidel outside the submitted work. Dr. Khan reports grants from United Therapeutics, grants from Johnson & Johnson, grants from 4D Medical, grants from Lung LLC, grants from Reata Pharmaceuticals, outside the submitted work. Dr. Kwon reports grants from CDC, during the conduct of the study. Dr. Lauring reports personal fees from Sanofi, personal fees from Roche, outside the submitted work. Dr. Lindsell reports grants from CDC, during the conduct of the study; grants from NIH, grants from DoD, grants from Marcus Foundation, other from bioMerieux, other from Endpoint LLC, other from Entegrion Inc, outside the submitted work; in addition, Dr. Lindsell has a patent for risk stratification in sepsis and septic shock issued. Dr. Martin reports grants from Vanderbilt University / Centers for Disease Control and Prevention, during the conduct of the study; personal fees from Pfizer, grants from Merck, outside the submitted work. Dr Monto reports consulting fees from Sanofi-Pasteur and Seqirus outside the submitted work. Dr. Peltan reports grants from Centers for Disease Control and Prevention, during the conduct of the study; grants from National Institutes of Health, grants from Janssen Pharmaceuticals, other from Asahi Kasei Pharma, other from Regeneron, outside the submitted work. Dr. Rice reports grants from Centers for Disease Control, during the conduct of the study; personal fees from Cumberland Pharmaceuticals, Inc, personal fees from Avisa Pharma, LLC, personal fees from Sanofi, outside the submitted work. Dr. Self reported research funding from CDC for the current project and consulting fees outside the submitted work from Aeprio Pharmaceuticals and Merck. The other authors reported no potential conflict of interest.

## Notes

**Funding:** Primary funding for this study was provided by the Centers for Disease Control and Prevention (75D30121F00002). The REDCap data tool was supported by a Clinical and Translational Science Award (UL1 TR002243) from the National Center for Advancing Translational Sciences.

### Clinical Trial

This is an observational study and not an interventional trial.

### Author Declarations

The following two Institutional Review Boards provided ethical approval for this work as a public health surveillance program with research exemption: [1] Vanderbilt University Medical Center (Nashville, Tennessee, United States; protocol number: 210357; determination: non-research public health surveillance; date of determination: 22 February 2021); and [2] The United States Centers for Disease Control and Prevention (Atlanta, Georgia, United States; protocol number: 0900f3eb81c1be34; determination: non-research public health surveillance; date of determination: 26 February 2021).

