## Supplemental Materials for "Effectiveness of SARS-CoV-2 mRNA Vaccines for Preventing Covid-19 Hospitalizations in the United States"

**Network:** The IVY Network

**Corresponding Author:** Wesley H. Self, MD, MPH; Vanderbilt University Medical Center; 312 Oxford House, 1313 21st Avenue South, Nashville, Tennessee 37232.; phone: 615-936-8047; fax: 615-936-3754.

**Funding:** Primary funding for this study was provided by the Centers for Disease Control and Prevention (75D30121F00002). The REDCap data tool was supported by a Clinical and Translational Science Award (UL1 TR002243) from the National Center for Advancing Translational Sciences.

**Disclaimer:** The findings and conclusions in this report are those of the authors and do not necessarily represent the official position of the Centers for Disease Control and Prevention.

#### Contents of Supplementary Material

**I. Supplementary Appendix A.** Investigators and Collaborators

Investigators and collaborators in the Influenza and Other Viruses in the Acutely Ill (IVY) Network

**Baylor, Scott and White, Temple, Texas**
Manjusha Gaglani, Tresa McNeal, Shekhar Ghamande, Nicole Calhoun, Kempapura Murthy, Judy Herrick, Amanda McKillop, Eric Hoffman, Martha Zayed, Michael Smith

**Baystate Medical Center, Springfield, Massachusetts**
Jay Steingrub, Ryan Kindle, Lori-Ann Kozikowski, Lesley De Souza, Scott Ouellette, Sherell Thornton-Thompson

**Beth Israel Medical Center, Boston Massachusetts**
Nate Shapiro, Patrick Tyler

**Centers for Disease Control and Prevention (CDC), Atlanta, Georgia**
Miwako Kobayashi, Samantha Olson, Manish Patel, Mark Tenforde, Meagan Stephenson, Stephanie Schrag, Jennifer Verani

**Cleveland Clinic, Cleveland, Ohio**
Abhijit Duggal, Omar Mehkri, Kiran Ashok, Susan Gole, Alexander King, Bryan Poynter

**Emory University, Atlanta, Georgia**
Laurence Busse, Caitlin ten Lohuis, Nicholas Stanley

**Hennepin County Medical Center, Minneapolis, Minnesota**
Matthew Prekker, Heidi Erickson, Audrey Hendrickson, Ellen Maruggi, Tyler Scharber, Leah Stodieck

**Intermountain Medical Center, Murray, Utah**
Ithan Peltan, Samuel Brown, Jeffrey Jorgensen, Robert Bowers, Jennifer King, Valerie Aston

**Johns Hopkins University, Baltimore, Maryland**
David N, Hager, Arber Shehu, Richard E. Rothman

**Montefiore Medical Center, Bronx, New York**
Michelle Gong, Amira Mohamed, Rahul Nair, Jen-Ting (Tina) Chen

**Ohio State Medical Center, Columbus, Ohio**
Matthew Exline, Sarah Karow, Emily Robart, Paulo Nunes Maldonado, Maryiam Khan, Preston So

**Oregon Health and Sciences University, Portland, Oregon**
Akram Khan, C. Terri Hough

**Stanford University, Stanford, California**
Jennifer G. Wilson, Alexandra June Gordon, Joe Levitt, Cynthia Perez, Anita Visweswaran, Jonasel Roque

**University of California-Los Angeles, Los Angeles, California**
Nida Qadir, Steven Chang, Adreanne Rivera, Trevor Frankel

**University of Colorado, Aurora, Colorado**
Adit Ginde, Josh Douin, Michelle Howell, Jennifer Friedel, Jennifer Goff, David Huynh, Michael Tozier, Conner Driver, Michael Carricato, Alexandra Foster

**University of Iowa, Iowa City, Iowa**
Nick Mohr, Anne Zepeski, Paul Nassar, Lori Stout, Zita Sibenaller, Alicia Walter, Jasmine Mares, Logan Olson, Bradley Clinansmith

**University of Miami, Miami, Florida**
Chris Mallow, Hayley Gershengorn, Carolina Rivas

**University of Michigan, Ann Arbor, Michigan**
Emily Martin, Arnold Monto, Adam Lauring, EJ McSpadden, Rachel Truscon, Anne Kaniclides, Lara Thomas, Ramsay Bielak, Weronika Damek Valvano, Rebecca Fong, William J. Fitzsimmons, Christopher Blair, Andrew L. Valesano, Julie Gilbert

**University of Washington, Seattle, Washington**
Daniel J. Henning, Christine D. Crider, Kyle A. Steinbock, Thomas C. Paulson, Layla A. Anderson

**Vanderbilt University Medical Center, Nashville, Tennessee**
Wesley H. Self, H. Keipp Talbot, Chris Lindsell, Carlos Grijalva, Ian Jones, Natasha Halasa, James Chappell, Kelsey Womack, Jillian Rhoads, Adrienne Baughman, Christy Kampe, Jakea Johnson, Rendie McHenry, Marcia Blair, Kim Hart, Robert McClellan, Todd Rice, Jonathan Casey, William B. Stubblefield, Douglas Conway

**Wake Forest University, Winston-Salem, North Carolina**
D. Clark Files, Kevin Gibbs, Mary LaRose, Leigha Landreth, Madeline Hicks, Lisa Parks

**Washington University, St. Louis, Missouri**
Hilary Babcock, Jennie Kwon, Jahnavi Bongu, David McDonald, Candice Cass, Sondra Seiler, David Park, Tiffany Hink, Meghan Wallace, Carey-Ann Burnham, Olivia G. Arter

### II. Supplementary Appendix B. Supplementary methods

#### 1. Eligibility criteria for participation

*Covid-19 Cases*

Summary for Cohort 1: Adult admitted to the hospital for acute Covid-19 who has tested positive for SARS-CoV-2.

Inclusion for Cohort 1 (cases):

1. Age ≥18 years old.
2. Hospital admission or in an emergency department awaiting hospital admission.
3. Symptoms and/or signs believed to be due to Covid-19, including at least 1 of the following: fever; cough; shortness of breath; loss of taste; loss of smell; use of respiratory support (high flow oxygen by nasal cannula, non-invasive ventilation or invasive ventilation) for the acute illness; new pulmonary findings on chest imaging consistent with pneumonia.
4. Clinically obtained test that is **positive** for acute SARS-CoV-2 infection. The positive test may be obtained before or after hospital arrival. Examples of acute SARS-CoV-2 tests include RT-PCR tests, nucleic acid amplification tests (NAAT), and antigen tests. Serology testing may not be used for eligibility.

Exclusion for Cohort 1 (cases):

1. Previous inclusion as a case.
2. The first positive test for acute SARS-CoV-2 infection is known to have occurred more than 10 days after onset of Covid-19 symptoms/signs listed in inclusion criterion #3. Patients with unknown onset date for Covid-19 symptoms/signs may be included.
3. Hospital presentation for the Covid-19 admission is known to have occurred more than 14 days after onset of Covid-19 symptoms/signs listed in inclusion criterion #3. Patients with unknown onset date for Covid-19 symptoms/signs may be included. Patients transferred from other hospitals may be included; the time of hospital presentation is the time of presentation to the first hospital.

*Test Negative Controls*

Summary for Cohort 2: Adult admitted to the hospital for an acute illness with symptom overlap with Covid-19 who has tested negative for SARS-CoV-2.

Inclusion for Cohort 2 (test negative controls):

1. Age ≥18 years old.
2. Hospital admission or in an emergency department awaiting hospital admission.
3. Symptoms and/or signs that overlap with Covid-19, including at least one of the following: fever; cough; shortness of breath; loss of taste; loss of smell; use of respiratory support (high flow oxygen by nasal cannula, non-invasive

ventilation or invasive ventilation) for the acute illness; new pulmonary findings on chest imaging consistent with pneumonia.

4. Clinically obtained test that is **negative** for acute SARS-CoV-2. The negative test may be obtained before or after hospital arrival. Examples of acute SARS-CoV-2 tests include RT-PCR tests, NAAT, and antigen tests. Serology testing may not be used for eligibility.

Exclusion for Cohort 2 (test negative controls):

1. Previous inclusion as a control.
2. The first negative test for acute SARS-CoV-2 infection is known to have occurred more than 10 days after onset of symptoms/signs listed in inclusion criterion #3. Patients with unknown onset date for Covid-19 symptoms/signs may be included.
3. Hospital presentation for the admission is known to have occurred more than 14 days after onset of symptoms/signs listed in inclusion criterion #3. Patients with unknown onset date for symptoms/signs may be included. Patients transferred from other hospitals may be included; the time of hospital presentation is the time of presentation to the first hospital.
4. Any positive test for acute SARS-CoV-2 infection in the 14 days prior to hospital presentation or between hospital presentation and inclusion (patients with a positive acute SARS-CoV-2 test should be screened for potential inclusion as a case).

*Syndrome-negative controls*

Summary for Cohort 3: Adult admitted to the hospital for a reason other than an acute respiratory illness and who does not have a clinical suspicion for Covid-19.

Inclusion for Cohort 3 (syndrome negative controls):

1. Age ≥18 years old.
2. Hospital admission or in an emergency department awaiting admission.
3. Clinical impression that Covid-19 is not the reason for admission.
4. None of the following signs or symptoms that overlap with Covid-19 in the past 14 days: fever; cough; shortness of breath; loss of taste; loss of smell; use of respiratory support (high flow oxygen by nasal cannula, non-invasive ventilation or invasive ventilation) for the acute illness; new pulmonary findings on chest imaging consistent with pneumonia.

Exclusion for Cohort 3 (syndrome negative controls):

1. Previous inclusion as a control.
2. Any positive test for acute SARS-CoV-2 infection in the 14 days prior to hospital presentation or between hospital presentation and inclusion (patients with a positive acute SARS-CoV-2 test should be screened for potential inclusion as a case).

**2. Model building for adjusted vaccine effectiveness estimates**

Vaccine effectiveness was determined by comparing the odds of antecedent SARS-CoV-2 vaccination in case-patients versus control participants. Several potential confounders were included as prespecified covariates in adjusted models of vaccine effectiveness in the base model:

1. Calendar time based on the date of admission in biweekly intervals using Morbidity and Mortality Weekly Report week surveillance cutoffs
2. U.S. Department of Health and Human Services region
3. Age in years as a continuous variable
4. Race and ethnicity specified in categories of non-Hispanic white, non-Hispanic black, Hispanic of any race, non-Hispanic other, or unknown/missing. Race and ethnicity were obtained by self-report
5. Female sex

Additional covariates were considered as potential confounders associated with likelihood of receiving a SARS-CoV-2 vaccine as well as risk of severe Covid-19 illness. A change in adjusted odds ratio from the base model of more than 5% in either direction was used as a prespecified cutoff for inclusion in final adjusted models. The following variables were considered:

1. ≥1 self-reported prior hospitalization in the previous year
2. Self-reported smoking, defined as current use or quitting less than 6 months ago
3. Number of chronic medical condition categories (cardiovascular disease, neurologic disease, pulmonary disease, gastrointestinal disease, endocrine disease, renal disease, hematologic disease, active malignancy, other immunosuppressive category, autoimmune condition) categorized as 0, 1, 2, or 3 or more
4. Number of medications the patient was taking as an outpatient prior the hospitalization as a continuous variable
5. Residence in a long-term care facility prior to hospitalization, which included nursing home, assisted living home, or rehab hospital / other sub-acute or chronic facility
6. Self-reported education level categorized as 1 or more years of college education versus high school graduation / GED or less
7. Self-reported mask use as "always" versus less than always based on response to the question: "In the 14 days prior to becoming ill, when you have gone outside of your home and been around other people (within 6 feet), how often did you wear a face covering (such as a cloth mask, surgical mask, dust mask or other respiratory, like an N95) that covered your nose and month?"
8. Self-reported attendance of an indoor event or gathering of more than 10 people as at least once versus never based on response to the question: "In the 14 days prior to becoming ill, how many days did you attend an indoor event or gathering with MORE than 10 people? This includes activities like attending church, parties, and sporting events, or visiting a bar or restaurant. This does not include shopping or errands."
9. Having medical insurance coverage versus being uninsured
10. Centers for Disease Control and Agency for Toxic Substances and Disease Registry Social Vulnerability Index (SVI) which uses 15 indicators grouped into 4 themes (socioeconomic status, household composition & disability, minority status and language, housing type & transportation) to generate an overall 0-1 composite ranking. National rankings were used for the analysis and SVI was modeled as a continuous variable. When patient home address was available, Census tract-level SVI was used. When full patient address was unavailable, Census tract-level SVI using the address of the admitting hospital was used.

**3. Definitions of underlying medical conditions**
Medical condition categories were defined using standardized definitions and obtained through electronic medical record (EMR) review by trained surveillance personnel.

Height and weight measurements were used to calculate obesity, defined as body-mass index ≥30 kg/$m^2$. Underlying condition categories evaluated in vaccine effectiveness models were defined as having documentation of 1 or more of the following by EMR review:

Chronic cardiovascular disease: Heart failure, peripheral vascular disease that limits mobility, prior myocardial infarction, cardiac arrhythmias including atrial fibrillation and ventricular arrhythmias, vascular heart disease, or hypertension

Chronic lung disease: Asthma, chronic obstructive pulmonary disease cystic fibrosis, pulmonary fibrosis, pulmonary hypertension, home oxygen use (except at night for sleep disorder), tracheostomy, home non-invasive ventilation use (except at night for sleep disorder), home invasive mechanical ventilation

Diabetes mellitus: Diabetes mellitus without end organ damage, diabetes mellitus with end organ damage

Immunocompromising condition: Active solid organ cancer without metastases (active cancer defined as treatment for the cancer or newly diagnosed cancer in the past 6 months), active solid organ cancer without metastases (active cancer defined as treatment for the cancer or newly diagnosed cancer in the past 6 months), active hematologic cancer (such as leukemia / lymphoma / myeloma) or active cancer defined as treatment for the cancer or newly diagnosed cancer in the past 6 months, human immunodeficiency virus (HIV) infection without AIDS, AIDS, congenital immunodeficiency syndrome, prior splenectomy, prior solid organ transplant, immunosuppressive medication, systemic lupus erythematosus, rheumatoid arthritis, psoriasis, scleroderma, inflammatory bowel disease including Crohn's disease or ulcerative colitis

**4. RT-PCR testing**
Upper respiratory swabs collected in viral transport medium and saliva were subjected to nucleic acid extraction and testing for SARS-CoV-2 at Vanderbilt University Medical Center according to the CDC EUA protocol, *CDC 2019-Novel Coronavirus (2019-nCoV) Real-Time RT-PCR Diagnostic Panel* (https://www.fda.gov/media/134922/download). Total nucleic acid was purified using the QiaCube HT (Qiagen) automated extraction system and QIAamp 96 Virus QiaCube HT kit and analyzed by RT-qPCR for SARS-CoV-2 nucleocapsid (N)-gene N1 and N2 targets and the human RNAse P (RNP) gene using StepOnePlus and QuantStudio 3 real-time PCR systems (Applied Biosystems). RNP served an endogenous indicator of specimen adequacy and sensor for PCR inhibitors. The pattern of N1, N2, and RNP Ct values served as a basis to ***1)*** assign a qualitative result of *positive* (N1 Ct <40, N2 Ct <40) or *not detected* (RNP Ct <40, N1 Ct ≥40, N2 Ct ≥40) or ***2)*** perform retesting using the original or freshly extracted nucleic acid template due to an inconclusive (N1 or N2 Ct <40 and the other target ≥40) or invalid (RNP, N1, and N2 Ct values each ≥40) result. Retested specimens were assigned a result of *positive*, *not detected*, *inconclusive*, or *invalid* based outcomes of secondary testing.

**5. Viral sequencing**
Specimen aliquots with positive RT-qPCR for either the N1 or N2 target with a cycle threshold ≤32 at the Vanderbilt University Medical Center laboratory were shipped to the University of Michigan on dry ice. RNA was extracted from 200µl transport media with the Thermo Fisher MagMAX Viral RNA Isolation Kit on a KingFisher instrument and eluted in

a 50µl volume. Extracted RNA was reverse transcribed with SuperScript IV (Thermo Fisher). For each sample, 1 µl of random hexamers and 1 µl of 10 mM dNTP were added to 11 µl of RNA, heated at 65˚C for 5 min, and placed on ice for 1 min. Then a reverse transcription master mix was added (4 µl of SuperScript IV buffer, 1 µl of 0.1M DTT, 1 µl of RNaseOUT RNase inhibitor, and 1 µl of SSIV reverse transcriptase) and incubated at 42˚C for 50 min, 70˚C for 10 min. Viral cDNA was amplified in two multiplex PCR reactions with the ARTIC Network version 3 primer pools and protocol using the Q5 Hot Start High-Fidelity DNA Polymerase (NEB) with the following thermocycler protocol: 98˚C for 30 s, then 35 cycles of 98˚C for 15 s, 63˚C for 5 min. Reaction products for a given sample were pooled together in equal volumes.

Libraries were prepared for sequencing with the Oxford Nanopore Technologies MinION using the ARTIC Network version 3 protocol. Samples were prepared in batches with one-pot native barcoding. Pooled PCR products were diluted in nuclease-free water with a dilution factor of 10. Amplicon ends were prepared for ligation with the NEBNext Ultra II End Repair/dA-Tailing Module (NEB). Unique barcodes (Oxford Nanopore Native Barcoding Expansion kits) were ligated per sample with the NEB Blunt/TA Ligase Master Mix. After barcoding, reactions were pooled together in equal volumes and purified barcoded amplicons with 0.4X volume of AMPure beads. Oxford Nanopore sequencing adapters were ligated with the NEBNext Quick Ligation Module (NEB) and the library was purified with 1X volume of AMPure beads. Final libraries were quantified with the Qubit 1X dsDNA HS Assay Kit (Thermo Fisher). Each library (15-20 ng) was loaded onto a flow cell (FLO-MIN106) and sequenced with a MinION.

Sequencing progress was monitored with RAMPART. Basecalling was performed with Guppy v4.0.14 and consensus genomes were called using the ARTIC Network bioinformatics pipeline (https://github.com/artic-network). PANGO lineage was assigned on genomes with >80% coverage using Pangolin v2.4.1 (https://pangolin.cog-uk.io, citation in main text). Genomes with >90% coverage were uploaded to GISAID (https://www.gisaid.org/).

**6. Sensitivity analyses**

Three sensitivity analyses were conducted to test key design features:

| Topic | Approach in Primary Analysis | Sensitivity Analysis |
|---|---|---|
| Source of SARS-CoV-2 vaccine information | Participates were considered vaccinated if they had source verification of vaccine receipt or self-report of a plausible date and location of vaccine receipt. | Participants with self-report vaccination without source verification were excluded. |
| Reference date to determine vaccination status for syndrome-negative control group | The reference date for the syndrome negative control group was 14 days prior to hospital admission. | The reference date for the syndrome negative control group was the median number of days between illness onset and admission for the test-negative control group plus 14 days. This more closely aligned the methods for determining the reference date for the syndrome negative control group and the case group. |
| Unknown date for onset of symptoms | Participants in the case and test-negative control groups with an unknown date for symptom onset had a symptom onset date assigned based on the median number of days between illness onset and admission date for patients with complete data within the same group. | Participants in the case and test-negative control group with an unknown date of illness onset were excluded. |

### III. Supplemental Tables

**Table S1.** Diagnoses for syndrome negative controls
Primary diagnosis for hospital admission for patients in the syndrome-negative control group (N=286) — IVY Network, United States, March–May 2021

| Reason | Number (%) |
|---|---:|
| Elective surgery | 16 (5.6) |
| Trauma | 20 (7.0) |
| Infection (not Covid-19) | 21 (7.3) |
| Cardiovascular illness | 31 (10.8) |
| Pulmonary illness | 2 (0.7) |
| Neurological illness | 18 (6.3) |
| Endocrine illness | 9 (3.2) |
| Genitourinary/renal illness | 13 (4.6) |
| Gastrointestinal illness | 50 (17.5) |
| Cancer related illness | 11 (3.9) |
| Other diagnosis | 95 (33.2) |

*Syndrome-negative control patients did not have Covid-19-like illness and tested negative for SARS-CoV-2.

**Table S2.** Sensitivity analyses
Sensitivity analyses for vaccine effectiveness of SARS-CoV-2 mRNA vaccines against hospitalized Covid-19— IVY Network, United States, March–May 2021

| Subgroup | No. of Vaccinated Case Patients / Total No. (%) | No. of Vaccinated Control Patients / Total No. (%) | Vaccine Effectiveness (95% Confidence Interval) |
|---|---|---|---|
| **Full vaccination** | | | |
| *Primary vaccine effectiveness model (reported in main text)* | 45/499 (9.0) | 215/489 (44.0) | 86.9 (80.4 to 91.2) |
| Verified vaccination only | 45/499 (9.0) | 206/480 (42.9) | 86.1 (79.3 to 90.7) |
| Imputing earlier reference date for syndrome-negative control group | 45/499 (9.0) | 208/482 (43.2) | 87.0 (80.6 to 91.3) |
| Excluding cases and test-negative controls without symptom onset date | 43/470 (9.2) | 204/459 (44.4) | 87.3 (80.7 to 91.6) |

* The analysis included case patients with Covid-19-like illness who tested positive for SARS-CoV-2 infection and control patients combined from two groups, including a test-negative control with Covid-19-like illness and syndrome-negative control without Covid-19-like illness, who tested negative for SARS-CoV-2. Vaccine effectiveness models were adjusted for calendar time in biweekly intervals, US Department of Health and Human Services region, age in years, sex, race and ethnicity. Vaccination status was classified based on the number of mRNA vaccine doses received before a reference date, which was defined as the date of symptom onset for cases and test-negative controls and date of hospital admission for syndrome-negative controls. Unvaccinated patients received no doses of vaccine before the reference date and fully vaccinated patients received both doses of vaccine ≥14 days before the reference date.

### IV. Supplemental Figures

**Figure S1.** Flow diagram of participants in the analysis

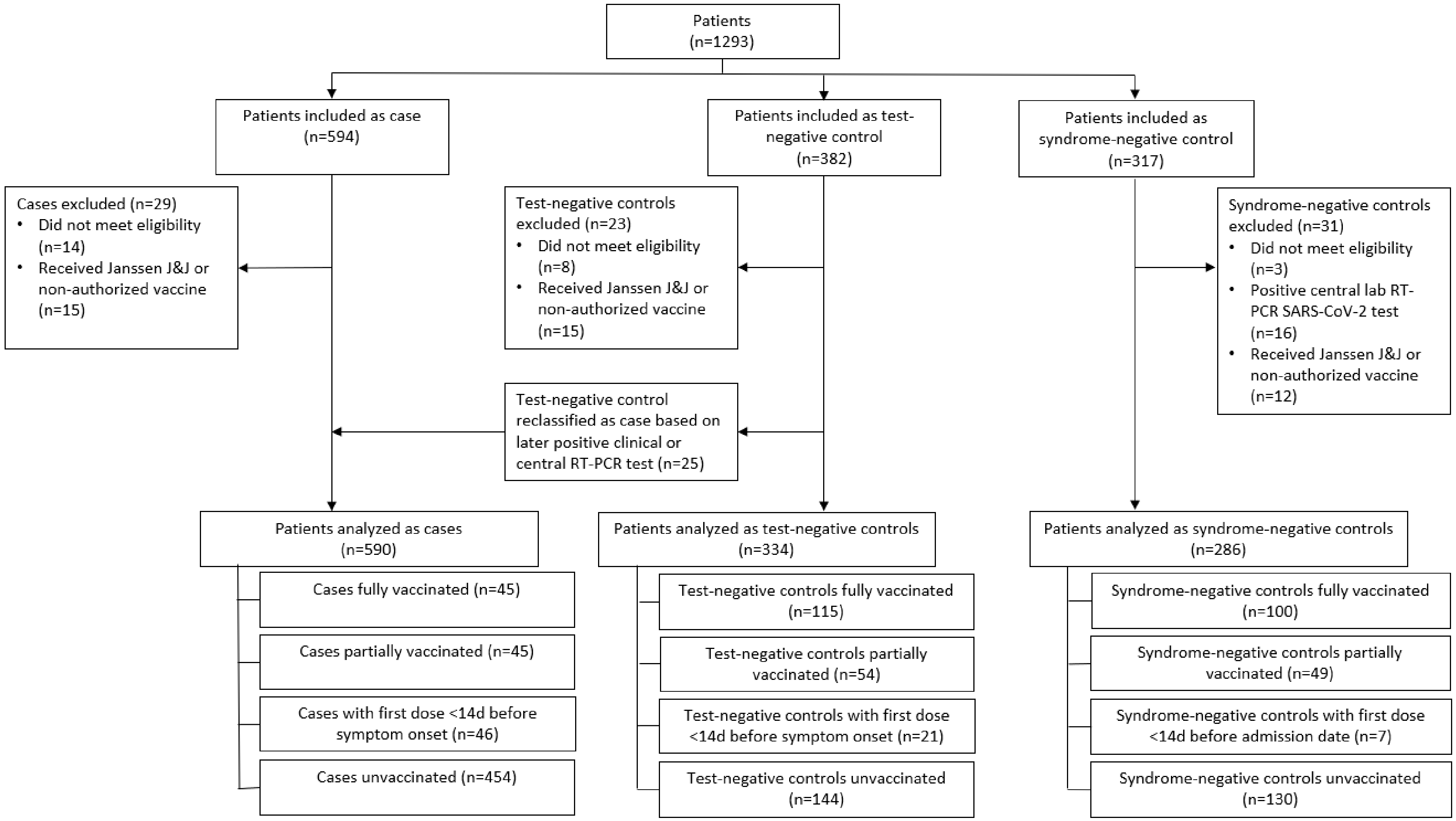

**Figure S2.** Participants by week.
Covid-19 cases and controls by week of hospital admission— IVY Network, United States, March–May 2021

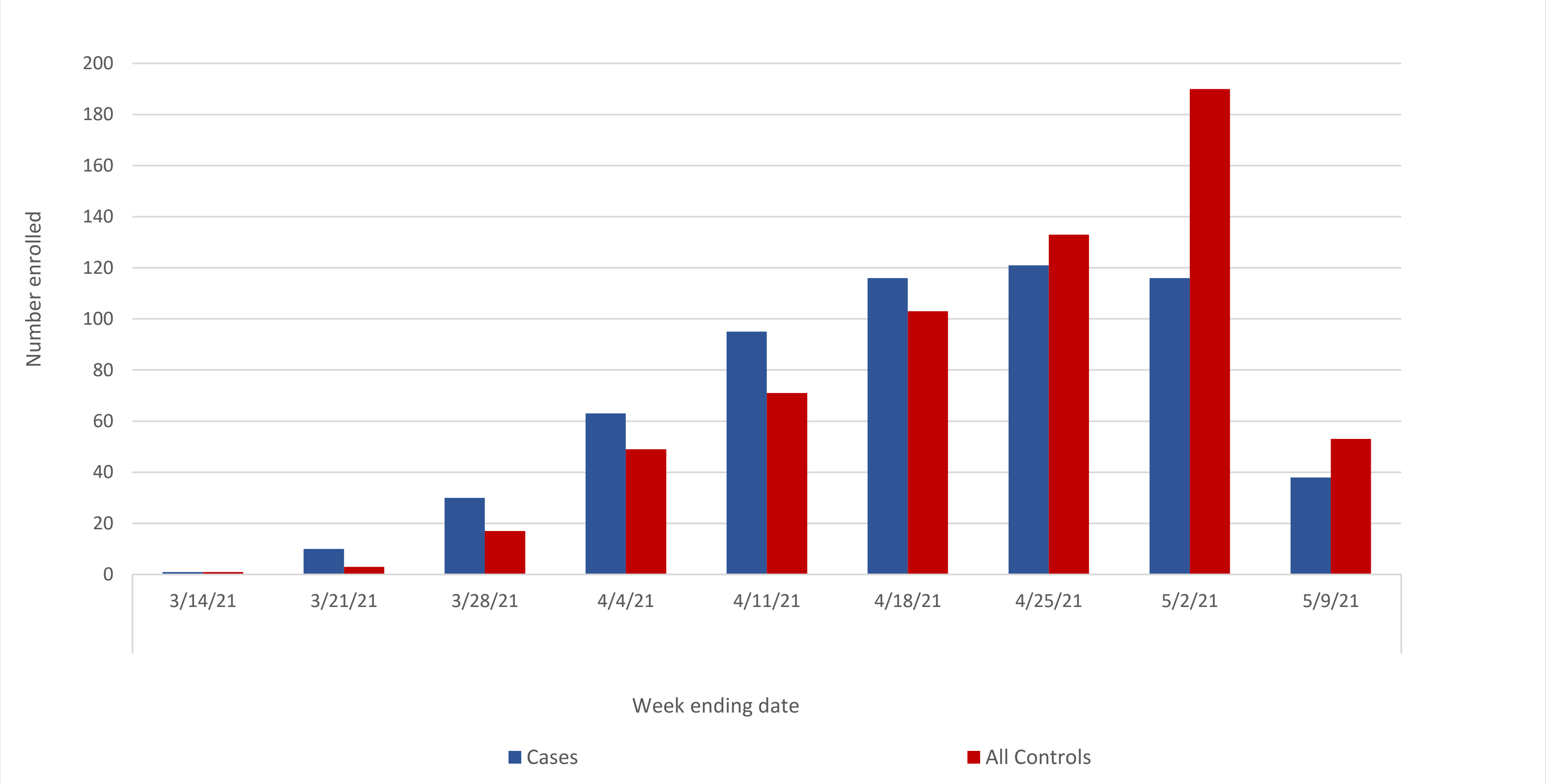

* The analysis included case patients with Covid-19-like illness who tested positive for SARS-CoV-2 infection and control patients combined from two groups, including a test-negative control with Covid-19-like illness and syndrome-negative control without Covid-19-like illness, who tested negative for SARS-CoV-2.

**Figure S3.** Vaccine effectiveness with test-negative control group only

Vaccine effectiveness of SARS-CoV-2 mRNA vaccines against hospitalized Covid-19 overall and by subgroup restricted to test-negative controls — IVY Network, United States, March–May 2021*

| Subgroup | Vaccinated case patients / total case patients (%) | Vaccinated control patients / total control patients (%) | Vaccine effectiveness (95% confidence interval) |
|---|---|---|---|
| **Full vaccination** | | | |
| Overall | 45/499 (9.0) | 115/259 (44.4) | 86.4 (78.7 to 91.3) |
| 18 to 49 yr | 1/186 (0.5) | 13/69 (18.8) | 97.6 (80.2 to 99.7) |
| 50 to 64 yr | 14/185 (7.6) | 21/74 (28.4) | 75.2 (41.2 to 89.5) |
| ≥65 yr | 30/128 (23.4) | 81/116 (69.8) | 88.7 (77.9 to 94.2) |
| Covid-19 vaccine product | | | |
| Pfizer-BioNTech | 28/482 (5.8) | 69/213 (32.4) | 84.5 (73.8 to 90.9) |
| Moderna | 17/471 (3.6) | 46/190 (24.2) | 88.7 (78.5 to 94.1) |
| B.1.1.7 variant lineage | 7/116 (6.0) | 115/259 (44.4) | 92.6 (82.0 to 96.9) |
| Underlying medical condition | | | |
| Chronic cardiovascular disease | 38/263 (14.5) | 83/181 (45.9) | 81.0 (67.8 to 88.8) |
| No chronic cardiovascular disease | 7/236 (3.0) | 32/78 (41.0) | 96.6 (90.2 to 98.8) |
| Chronic lung disease | 14/98 (14.3) | 39/94 (41.5) | 80.8 (54.8 to 91.9) |
| No chronic lung disease | 31/398 (7.8) | 76/165 (46.1) | 89.7 (81.9 to 94.2) |
| Diabetes mellitus | 19/151 (12.6) | 42/98 (42.9) | 78.3 (54.1 to 89.7) |
| No diabetes mellitus | 26/347 (7.5) | 73/161 (45.3) | 90.3 (82.4 to 94.7) |
| Immunocompromising condition | 20/79 (25.3) | 31/68 (45.6) | 45.7 (-34.3 to 78.1) |
| No immunocompromising condition | 25/417 (6.0) | 84/191 (44.0) | 91.7 (85.5 to 95.3) |
| Obesity | 21/289 (7.3) | 54/122 (44.3) | 88.5 (78.0 to 94.0) |
| No obesity | 24/203 (11.8) | 61/137 (44.5) | 82.3 (66.1 to 90.7) |
| **Partial vaccination** | | | |
| Overall | 45/499 (9.0) | 54/198 (27.3) | 75.2 (60.2 to 84.6) |

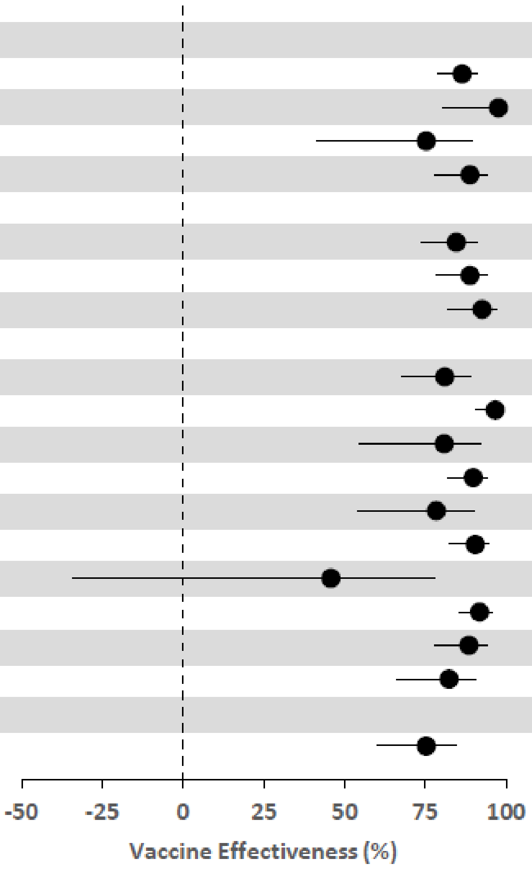

* The analysis included case patients with Covid-19-like illness who tested positive for SARS-CoV-2 infection and test-negative controls with Covid-19-like illness but who tested negative for SARS-CoV-2. Vaccine effectiveness models were adjusted for calendar time in biweekly intervals, US Department of Health and Human Services region, age in years, sex, and race and ethnicity. Vaccination status was classified based on the number of vaccine doses received before a reference date, which was defined as the date of symptom onset for cases and test-negative controls and date of hospital admission for syndrome-negative controls. Unvaccinated patients received no doses of vaccine before the reference date, partially vaccinated patients received one of two doses of vaccine ≥14 days before the reference date or both doses with the second dose received <14 days before the reference date, and fully vaccinated patients received both doses of vaccine ≥14 days before the reference date.

**Figure S4**. Vaccine effectiveness with syndrome-negative control group only

Vaccine effectiveness of SARS-CoV-2 mRNA vaccines against hospitalized Covid-19 overall and by subgroup restricted to syndrome-negative controls — IVY Network, United States, March–May 2021*

| Subgroup | Vaccinated case patients / total case patients (%) | Vaccinated control patients / total control patients (%) | Vaccine effectiveness (95% confidence interval) |
|---|---|---|---|
| **Full vaccination** | | | |
| Overall | 45/499 (9.0) | 100/230 (43.5) | 86.8 (78.7 to 91.8) |
| 18 to 49 yr | 1/186 (0.5) | 12/71 (16.9) | 97.1 (74.0 to 99.7) |
| 50 to 64 yr | 14/185 (7.6) | 18/62 (29.0) | 81.5 (52.5 to 92.8) |
| ≥65 yr | 30/128 (23.4) | 70/97 (72.2) | 85.5 (71.7 to 92.6) |
| Covid-19 vaccine product | | | |
| Pfizer-BioNTech | 28/482 (5.8) | 53/183 (29.0) | 83.5 (70.7 to 90.7) |
| Moderna | 17/471 (3.6) | 47/177 (26.6) | 90.4 (81.1 to 95.1) |
| B.1.1.7 variant lineage | 7/116 (6.0) | 100/230 (43.5) | 93.7 (83.9 to 97.5) |
| Underlying medical condition | | | |
| Chronic cardiovascular disease | 38/263 (14.5) | 67/130 (51.5) | 85.9 (74.2 to 92.3) |
| No chronic cardiovascular disease | 7/236 (3.0) | 33/100 (33.0) | 93.1 (81.3 to 97.4) |
| Chronic lung disease | 14/98 (14.3) | 33/64 (51.6) | 86.1 (61.0 to 95.0) |
| No chronic lung disease | 31/398 (7.8) | 67/166 (40.4) | 87.2 (77.5 to 92.7) |
| Diabetes mellitus | 19/151 (12.6) | 31/64 (48.4) | 87.4 (70.1 to 94.7) |
| No diabetes mellitus | 26/347 (7.5) | 69/166 (41.6) | 88.0 (77.8 to 93.5) |
| Immunocompromising condition | 20/79 (25.3) | 25/48 (52.1) | 75.0 (27.5 to 91.4) |
| No immunocompromising condition | 25/417 (6.0) | 75/182 (41.2) | 90.7 (83.0 to 94.8) |
| Obesity | 21/289 (7.3) | 41/93 (44.1) | 89.4 (77.4 to 95.1) |
| No obesity | 24/203 (11.8) | 59/136 (43.4) | 84.4 (68.9 to 92.2) |
| **Partial vaccination** | | | |
| Overall | 45/499 (9.0) | 49/179 (27.4) | 79.2 (65.3 to 87.6) |

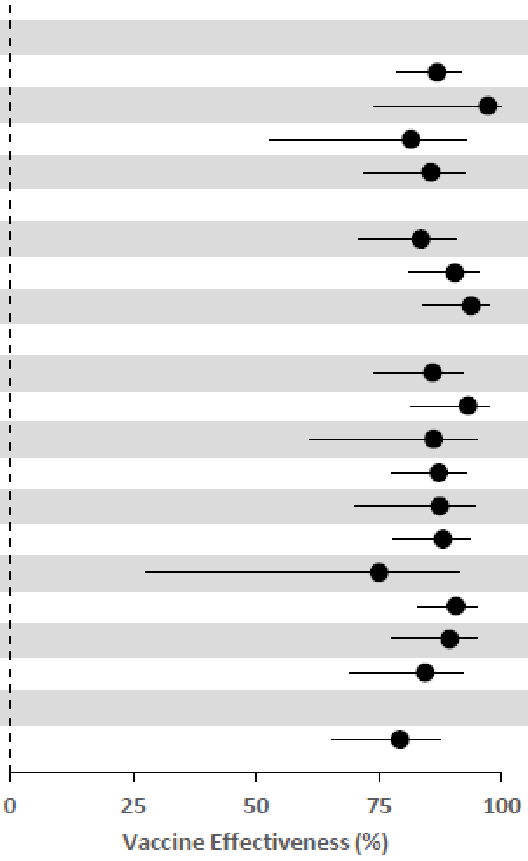

* The analysis included case patients with Covid-19-like illness who tested positive for SARS-CoV-2 infection and syndrome-negative control without Covid-19-like illness who tested negative for SARS-CoV-2. Vaccine effectiveness models were adjusted for calendar time in biweekly intervals, US Department of Health and Human Services region, age in years, sex, and race and ethnicity. Vaccination status was classified based on the number of vaccine doses received before a reference date, which was defined as the date of symptom onset for cases and test-negative controls and date of hospital admission for syndrome-negative controls. Unvaccinated patients received no doses of vaccine before the reference date, partially vaccinated patients received one of two doses of vaccine ≥14 days before the reference date or both doses with the second dose received <14 days before the reference date, and fully vaccinated patients received both doses of vaccine ≥14 days before the reference date.
