## Supplementary material for "Effectiveness of SARS-CoV-2 mRNA Vaccines for Preventing Covid-19 Hospitalizations in the United States": Protocol and Statistical Analysis Plan

This supplement contains:

1. Study protocol (19 pages)
2. Study statistical analysis plan, including a summary of changes between the original and final analysis plan (13 pages)

Corresponding Author:
Wesley H. Self, MD, MPH; Vanderbilt University Medical Center; 312 Oxford House, 1313 21st Avenue South, Nashville, Tennessee 37232.; phone: 615-936-8047; fax: 615-936-3754.

IVY

INFLUENZA AND OTHER VIRUSES IN THE ACUTELY ILL

Protocol

**Adult Inpatient SARS-CoV-2 Vaccine Effectiveness Surveillance**

| | |
|---|---|
| Full Title | Adult Inpatient SARS-CoV-2 Vaccine Effectiveness Surveillance |
| IVY protocol number | IVY-3 |
| Short title | IVY-3 SARS-CoV-2 VE |
| Protocol version | 1 |
| Date | February 16, 2021 |
| Funder | Centers for Disease Control and Prevention (CDC) |
| Network | Influenza and Other Viruses in the Acutely Ill (IVY) |
| Coordinating Center | Vanderbilt University Medical Center |
| Principal Investigator | Wesley H. Self, MD, MPH<br>Vanderbilt University Medical Center |
| CDC Scientists | Miwako Kobayashi, MD, MPH<br>Mark Tenforde, MD, PhD<br>Manish M. Patel, MD<br>Jennifer R. Verani, MD, MPH |

**IVY**

**INFLUENZA AND OTHER VIRUSES IN THE ACUTELY ILL**

| Participating Institutions and Protocol Committee | | | |
|---|---|---|---|
| **#** | **Institution** | **Location** | **Investigators** |
| 1 | Baylor, Scott & White | Temple, TX | Manjusha Gaglani, Tresa McNeal, Shekhar Ghamande |
| 2 | Baystate Medical Center | Springfield, MA | Jay Steingrub, Howard Smithline |
| 3 | Beth Israel Deaconess | Boston, MA | Nate Shapiro, Patrick Tyler |
| 4 | Cleveland Clinic | Cleveland, OH | Abhijit Duggal |
| 5 | Emory University | Atlanta, GA | Larry Busse |
| 6 | Hennepin County Medical Center | Minneapolis, MN | Matt Prekker, Heidi Erickson |
| 7 | Intermountain Medical Center | Murray, UT | Ithan Peltan, Sam Brown |
| 8 | Johns Hopkins | Baltimore, MD | David Hager, Jeremiah Hinson |
| 9 | Montefiore Medical Center | Bronx, NY | Michelle Gong, Amira Mohamed, Rahul Nair, Jen-Ting (Tina) Chen |
| 10 | Ohio State Medical Center | Columbus, OH | Matt Exline |
| 11 | Oregon Health & Sciences University | Portland, OR | Akram Khan, Terri Hough |
| 12 | Stanford University | Stanford, CA | Jenny Wilson, Joe Levitt |
| 13 | University of California, Los Angeles | Los Angeles, CA | Nida Qadir, Steve Chang |
| 14 | University of Colorado | Aurora, CO | Adit Ginde, Josh Douin |
| 15 | University of Iowa | Iowa City, IA | Nick Mohr, Anne Zepeski, Paul Nassar |
| 16 | University of Miami | Miami, FL | Chris Mallow, Hayley Gershengorn |
| 17 | University of Michigan | Ann Arbor, MI | Emily Martin, Arnold Monto, Adam Lauring |
| 18 | University of Washington | Seattle, WA | Dan Henning |
| 19 | Vanderbilt University Medical Center | Nashville, TN | Todd Rice, Jon Casey |
| 20 | Wake Forest University | Winston-Salem, NC | D. Clark Files, Kevin Gibbs |
| 21 | Washington University | St. Louis, MO | Hillary Babcock, Jennie Kwon |
| Coordinating Center (Vanderbilt University Medical Center) | | Nashville, TN | Wesley Self, Keipp Talbot, Chris Lindsell, Carlos Grijalva, Ian Jones, Natasha Halasa, Jim Chappell, Kelsey Womack, Jillian Rhoads, Adrienne Baughman, Christy Kampe |
| Centers for Disease Control and Prevention | | Atlanta, GA | Miwako Kobayashi, Manish M. Patel, Mark Tenforde, Jennifer R. Verani |

**Protocol: Table of Contents**

### 1. Summary of Protocol Changes

| | |
|---|---|
| Protocol Version: | 1 |
| Protocol Date: | February 16, 2021 |
| Summary of changes: | N/A, original protocol |

### 2. Summary of Protocol

Adult Inpatient SARS-CoV-2 Vaccine Effectiveness Surveillance is an observational, multicenter, prospective public health surveillance activity conducted by the Centers for Disease Control and Prevention (CDC) in collaboration with the *Influenza and other Viruses in the acutelY ill* (IVY) Network. We will enroll hospitalized adults with Coronavirus Disease 2019 (COVID-19) and laboratory-confirmed severe acute respiratory syndrome coronavirus 2 (SARS-CoV-2) infection. Additionally, we will also enroll two control groups: (1) adults hospitalized with signs and symptoms of an acute respiratory infection (ARI) who test negative for SARS-CoV-2 [test negative controls], and (2) adults hospitalized without signs and symptoms of an ARI [syndrome negative controls]. We will collect data on prior receipt of SARS-CoV-2 vaccination from each enrolled patient.

By comparing the odds of vaccination among enrolled cases and controls, we will calculate vaccine effectiveness for the prevention of COVID-19-associated hospitalizations. We will compare vaccine effectiveness of different types of SARS-CoV-2 vaccines. We will also estimate vaccine effectiveness for different variants of SARS-CoV-2 and by baseline patient characteristics, including age group, race/ethnicity, and comorbidity burden. We will collect an expanded set of biospecimens in a subset of enrolled patients to evaluate biological characteristics of SARS CoV-2 infection and SARS-CoV-2 vaccination, including the kinetics and durability of anti-SARS-CoV-2 antibodies, the respiratory tract microbiome and virome, and T and B cell responses.

Data collected in this surveillance project will be used to serially calculate SARS-CoV-2 vaccine effectiveness estimates in the United States and inform CDC public health policies.

### 3. Background

On March 11, 2020, the World Health Organization declared Coronavirus Disease 2019 (COVID-19) a pandemic. Through February 14, 2021, over 100 million confirmed cases and 2.3 million deaths from COVID-19 have been reported worldwide.[1] Multiple vaccines targeting severe acute respiratory syndrome coronavirus 2 (SARS-CoV-2), the virus that causes COVID-19, have demonstrated efficacy for preventing COVID-19 in phase III clinical trials.[2,3] Implementation of SARS-CoV-2 vaccination, initially in high-priority populations followed by the general public, is underway in the United States. Given the rapid pace of vaccine development and the critical role of vaccines to control the pandemic, data on the protective benefits under real-world conditions (vaccine effectiveness) are urgently needed.[4]

In this multicenter prospective surveillance project, the *Influenza and other Viruses in the acutelY ill* (IVY) Network investigators will evaluate SARS-CoV-2 vaccine effectiveness for the prevention of COVID-19 hospitalizations during implementation of the SARS-CoV-2 vaccination program in the United States. Key assessments will include the duration of protection from vaccines, protection against different SARS-CoV-2 variants, comparative effectiveness of different vaccine types, effectiveness in different subgroups of the population, and mechanisms of vaccine failures.

### 4. Objectives

1. Estimate vaccine effectiveness for the SARS-CoV-2 vaccines in use in the United States to prevent COVID-19 hospitalization among adults.
2. Estimate SARS-CoV-2 vaccine effectiveness for the different SARS-CoV-2 vaccines in use in the United States for prevention of COVID-19 hospitalizations.
3. Estimate SARS-CoV-2 vaccine effectiveness against circulating SARS-CoV-2 variants for prevention of COVID-19 hospitalizations.
4. Estimate SARS-CoV-2 vaccine effectiveness for prevention of COVID-19 hospitalizations within subgroups of the population defined by age, race/ethnicity, and comorbidities.
5. Assess the duration of protection for SARS-CoV-2 vaccination.
6. Assess the clinical validity of SARS-CoV-2 reverse transcriptase polymerase chain reaction (RT-PCR) results.
7. Obtain biological specimens to study the biology of severe COVID-19, the effects of vaccination, and mechanisms of vaccine failure.

### 5. Schedule of Events

The schedule of events for this program is outlined in Table 1.

**Table 1**. Schedule of events.

| **Activity** | **Enrollment (Day 1)** | **Day 3** | **Day 7** | **End of hospitalization** | **Day 28** |
|---|---|---|---|---|---|
| Confirm eligibility | X | | | | |
| Patient/surrogate interview | X | | | | |
| Nasal or saliva specimen collection | X | | | | X* |
| Medical record review | X | | | X | |
| Vaccine verification | X | | | | X |
| Two-clinician assessment for COVID-19 diagnosis (subset of patients) | | | | X* | |
| Blood specimen collection (subset of patients) | X* | X* | X* | X* | X* |

*Events only completed on a subset of patients at sites that agree to participate in expanded activities.

### 6. Population

#### 6.1 Approach to Enrollment

Patients will be enrolled in the hospital setting at sites affiliated with the IVY Network. Study personnel will screen medical records and patient logs to identify potentially eligible patients and prospectively approach those patients to confirm eligibility. Patients confirmed to be eligible will be enrolled at that time. Enrollment (study day 1) is defined as the calendar day study personnel confirmed eligibility for enrollment and collected the first element of data for the study. Study entry will be signified by assignment of a study ID number in REDCap.

#### 6.2. Eligibility Criteria

Three separate cohorts will be included in this study. Eligibility criteria vary by cohort and are detailed in this section.

#### *6.2.i. Cohort 1: SARS CoV-2 Cases*

Summary for Cohort 1: Adult admitted to the hospital for acute COVID-19 who has tested positive for SARS-CoV-2.

Inclusion for Cohort 1 (cases):

1. Age ≥18 years old.
2. Hospital admission or in an emergency department awaiting hospital admission.
3. Symptoms and/or signs believed to be due to COVID-19, including at least 1 of the following: fever; cough; shortness of breath; loss of taste; loss of smell; use of respiratory support (high flow oxygen by nasal cannula, non-invasive ventilation or invasive ventilation) for the acute illness; new pulmonary findings on chest imaging consistent with pneumonia.
4. Clinically obtained test that is **positive** for acute SARS-CoV-2 infection. The positive test may be obtained before or after hospital arrival. Examples of acute SARS-CoV-2 tests include RT-PCR tests, nucleic acid amplification tests (NAAT), and antigen tests. Serology testing may not be used for eligibility.

Exclusion for Cohort 1 (cases):

1. Previous enrollment as a case in this study.
2. The first positive test for acute SARS-CoV-2 infection is known to have occurred more than 10 days after onset of COVID-19 symptoms/signs listed in inclusion criterion #3. Patients with unknown onset date for COVID-19 symptoms/signs may be enrolled.
3. Hospital presentation for the COVID-19 admission is known to have occurred more than 14 days after onset of COVID-19 symptoms/signs listed in inclusion criterion #3. Patients with unknown onset date for COVID-19 symptoms/signs may be enrolled. Patients transferred from other hospitals may be enrolled; the time of hospital presentation is the time of presentation to the first hospital.

#### *6.2.ii. Cohort 2: SARS-CoV-2 Test Negative Controls*

Summary for Cohort 2: Adult admitted to the hospital for an acute illness with symptom overlap with COVID-19 who has tested negative for SARS-CoV-2.

Inclusion for Cohort 2 (test negative controls):

1. Age ≥18 years old.
2. Hospital admission or in an emergency department awaiting hospital admission.
3. Symptoms and/or signs that overlap with COVID-19, including at least 1 of the following: fever; cough; shortness of breath; loss of taste; loss of smell; use of respiratory support (high flow oxygen by nasal cannula, non-invasive ventilation or invasive ventilation) for the acute illness; new pulmonary findings on chest imaging consistent with pneumonia.
4. Clinically obtained test that is **negative** for acute SARS-CoV-2. The negative test may be obtained before or after hospital arrival. Examples of acute SARS-CoV-2 tests include RT-PCR tests, NAAT, and antigen tests. Serology testing may not be used for eligibility.

Exclusion for Cohort 2 (test negative controls):

1. Previous enrollment as a control in this study.
2. The first negative test for acute SARS-CoV-2 infection is known to have occurred more than 10 days after onset of symptoms/signs listed in inclusion criterion #3. Patients with unknown onset date for COVID-19 symptoms/signs may be enrolled.
3. Hospital presentation for the admission is known to have occurred more than 14 days after onset of symptoms/signs listed in inclusion criterion #3. Patients with unknown onset date for symptoms/signs may be enrolled. Patients transferred from other hospitals may be enrolled; the time of hospital presentation is the time of presentation to the first hospital.
4. Any positive test for acute SARS-CoV-2 infection in the 14 days prior to hospital presentation or between hospital presentation and enrollment (patients with a positive acute SARS-CoV-2 test should be screened for potential enrollment as a case).

##### *6.2.iii. Cohort 3: SARS-CoV-2 Syndrome Negative Controls*

Summary for Cohort 3: Adult admitted to the hospital for a reason other than an acute respiratory illness and who does not have a clinical suspicion for COVID-19.

Inclusion for Cohort 3 (syndrome negative controls):

1. Age ≥18 years old.
2. Hospital admission or in an emergency department awaiting admission.
3. Clinical impression that COVID-19 is not the reason for admission.
4. None of the following signs or symptoms that overlap with COVID-19 in the past 14 days: fever; cough; shortness of breath; loss of taste; loss of smell; use of respiratory support (high flow oxygen by nasal cannula, non-invasive ventilation or invasive ventilation) for the acute illness; new pulmonary findings on chest imaging consistent with pneumonia.

Exclusion for Cohort 3 (syndrome negative controls):

1. Previous enrollment as a control in this study.
2. Any positive test for acute SARS-CoV-2 infection in the 14 days prior to hospital presentation or between hospital presentation and enrollment (patients with a positive acute SARS-CoV-2 test should be screened for potential enrollment as a case).

#### 6.3 Case: Control Enrollment Ratios

Sites will enroll approximately equal numbers of cases, test-negative controls, and syndrome-negative controls (case: test-negative control: syndrome negative ratio = 1:1:1). Enrollment of each case will prompt a site to enroll a test-negative control and a syndrome-negative control. Enrollment ratios may be altered by CDC and the IVY protocol committee based on evolution of the COVID-19 pandemic.

### 7. Data Collection

#### 7.1 Patient Interview

At the time of enrollment, study personnel will conduct an interview with the patient and/or surrogates. Key data from the patient interview will include information on SARS-CoV-2 vaccination, patient-reported reasons for receiving or not receiving vaccination, general history of vaccine usage, demographics, history of healthcare utilization, acute symptoms, risk factors for severe infection, antiviral use, baseline health status, and medical comorbidities. If an enrolled patient is unable to provide answers to the interview questions, study personnel will seek to ask a surrogate the interview questions. If neither the patient nor a surrogate can answer interview questions, study personnel will answer as many of the interview questions as possible from medical chart review.

#### 7.2 Chart Review

Study personnel will complete a medical record chart review including data generated before and during the hospitalization in which the patient was enrolled, truncated at hospital day 28. The chart review will include information through the earlier of hospital discharge, death, or 28 days after initial hospital presentation for the hospitalization in which the patient was enrolled. Hence, the time frame for outcomes will be in-hospital 28-day outcomes. Key data collected by chart review will include: pre-admission home type (e.g. community, assisted living, nursing home), medical comorbidities; treatments received in the hospital; results of clinically-obtained SARS-CoV-2 tests, microbiology tests, and biomarker tests (e.g. lactate, procalcitonin, CRP); death; respiratory support; shock; vasopressor use; renal replacement therapy; extracorporeal membrane oxygenation (ECMO); length of stay; and discharge destination.

#### 7.3 Database

Data will be entered by personnel at each enrolling site into an electronic database housed within REDCap.[5] Centralized data management will be conducted at Vanderbilt; this work will include maintaining the electronic data tools, cleaning the dataset, issuing data queries, and working with sites to resolve data queries. Vanderbilt will house the final dataset and will share it with CDC and IVY investigators. The final dataset shared with investigators will not contain patient identifiers.

### 8. Viral Testing

#### 8.1 Respiratory Samples

Enrolled patients will have upper respiratory specimens collected for viral testing in the study’s central laboratory. The preferred method of upper respiratory specimen collection is by fresh nasal (mid-turbinate) swabbing by research personnel. Alternatively, a saliva sample may be collected or a residual respiratory sample collected clinically from earlier in the hospitalization may be collected from the clinical laboratory without fresh swabbing. For nasal swabs collected for the study, flocked swabs will be used to obtain a mid-turbinate sample. Samples will be placed in viral transport media, frozen, and shipped to the central laboratory for testing.

#### 8.2 Viral Laboratory

Respiratory samples collected at enrolling sites will be shipped to the central laboratory at Vanderbilt University Medical Center for viral testing. Each sample will be tested for SARS-CoV-2 with reverse transcription quantitative polymerase chain reaction (RT-qPCR). Additional viral targets will be determined by CDC based on circulating viruses and public health needs. Testing for the human RNAseP (RNP) gene will be performed as an internal positive control for human nucleic acid. If influenza testing is completed, influenza positive results will be further tested to determine A subtypes and B lineages (Victoria and Yamagata). Residual samples remaining after viral testing at Vanderbilt will be frozen and shipped to CDC.

### 9. Vaccine Verification

The primary exposure variable in SARS-CoV-2 vaccine effectiveness analyses will be receipt of a SARS-CoV-2 vaccine. Therefore, we will undertake intensive vaccine verification efforts including the following steps: 1) at the time of enrollment, obtain patient/surrogate report of SARS-CoV-2 vaccination and signed permission to search vaccination registries, medical records, and records from pharmacies, employers, and other non-traditional vaccination locations; 2) ask participants to view their “SARS-CoV-2 vaccine card,” which is a paper form used to document SARS-CoV-2 vaccine receipt by many vaccination centers; 3) systematic search of local electronic medical records; 4) search of state vaccination registry; 5) contact relevant pharmacies, clinics (e.g. primary care provider), payors and other venues for evidence of vaccination; 6) call patients/surrogates to clarify discordant information between initial self-/surrogate-report and results of systematic searches of medical records, registries, and vaccination venues.

For consistency, the verification process will encompass similar efforts to confirm absence of vaccination among those who self-reported no vaccination as to confirm the presence of vaccination among those who self-report vaccination. State registries will be reviewed twice: once during the vaccine verification for each patient at the time of enrollment and once at least 28 days later to capture any delays in data transfer to the registry. For each vaccination identified, we will collect vaccination date, vaccine product, and lot number.

### 10. Expanded Study Procedures

Expanded biospecimen and data collection will be completed for a subset of enrolled patients at sites that agree to participate in expanded study procedures.

#### 10.1 Expanded Biospecimen Collection

Samples outlined in Table 2 will be collected. The IVY coordinating center will assign the number of patients for sample collection at each site that agrees to participate in this component of the study. Samples will be collected at sites, temporarily stored locally, and then shipped to Vanderbilt University Medical Center for analysis and/or delivery to other laboratories for analysis.

The Day 1, 3, 7 and discharge (DC) samples will be collected if the patient remains in the hospital at those time points (that is, home visits will not be completed for these time points). Convalescent sera samples will be collected around Day 28. These convalescent sera sample may be collected in the hospital (if the patient remains hospitalized), or during a study visit after discharge. Study personnel may perform home visits to collect the Day 28 samples.

Approximately 400 participants (300 cases, 50 test-negative controls, 50 syndrome negative controls) will be selected for collection of acute sera, plasma, and nasal microbiome samples. Approximately 100 participants (all cases) will be selected for PBMC collection. A subset of approximately 250 participants (200 cases and 50 test negative controls) with acute sera collected will be selected for convalescent visit sample collection at Day 28. Samples collected during the convalescent visit will include sera, plasma, and a nasal swab or saliva for SARS-CoV-2 testing. Patients selected for convalescent sample collection at Day 28 will also undergo the expanded data collection for inclusion in the sub-study to assess clinical validity of SARS-CoV-2 RT-PCR results. Sample size targets for biospecimen collection may increase based on SARS-CoV-2 activity and evolution of the COVID-19 pandemic.

**Table 2.** Expanded biospecimen collection.

| Specimen type | Days of Collection* | Sample size goal across all sites | Purpose |
|---|---|---|---|
| Acute serum | 1, 3, 7, DC | 400<br>(300 cases, 50 test negative controls, 50 syndrome negative controls) | Kinetics and durability of antibody response, correlates of protection |
| Plasma | 1, 3, 7, DC | 400<br>(300 cases, 50 test negative controls, 50 syndrome negative controls) | Proteomics, biomarkers |
| Nasal microbiome sample | 1 | 400<br>(300 cases, 50 test negative controls, 50 syndrome negative controls) | Microbiome and virome |
| PBMCs | 1, 3, 7, DC | 100<br>(100 cases) | T and B cell responses |
| Convalescent visit samples:<br>serum<br>plasma<br>nasal swab or saliva | 28 | 250<br>(subset of patients with acute serum; 200 cases, 50 test negative controls) | Kinetics and durability of antibody response.<br>Biomarkers of inflammation.<br>SARS-CoV-2 RT-PCR. |

*Explanation for days of collection: Day 1 = enrollment day, as close to hospital presentation as possible; Day 3 = approximately 48 hours after enrollment (window: 24 hours – 72 hours after enrollment); Day 7 = approximately 168 hours after enrollment (window: 144 hours (6 days) – 192 hours (8 days) after enrollment; D/C = as close to discharge as possible for patients hospitalized over 10 days; Day 28 window for sample collection is between Day 28 and Day 42.

#### 10.2 Expanded Data Collection

The patients selected for convalescent visit (approximately 250 patients, including approximately 200 cases and 50 test negative controls) will undergo expanded data collection for a sub-study evaluating the clinical validity of SARS-CoV-2 RT-PCR tests. Each of these patients will undergo a chart review by two clinicians blinded to SARS-CoV-2 RT-PCR results. Using a standardized case report form, the clinicians will document their impression on whether the patient had COVID-19. These clinical impressions will be used as a comparison for SARS-CoV-2 RT-PCR test results (see analysis section).

### 11. Analysis

#### 11.1 Vaccine Effectiveness Analyses

##### *11.1.i Classification of case-control status*

Vaccine effectiveness will be calculated by comparing the odds of SARS-CoV-2 vaccination among cases and each of the control groups. Patients enrolled with COVID-like illness (enrollment cases and test negative controls) who test positive for SARS-CoV-2 by a clinical test or by the centralized research RT-PCR test will be classified as cases for the analysis ("analytical cases"). Patients enrolled with COVID-like illness who test negative for SARS-CoV-2 by all clinical and research testing (enrollment test-negative controls who do not have a positive RT-PCR for SARS-CoV-2 by centralized research testing) will be classified as test-negative controls for the analysis ("analytical test negative controls"). Patients enrolled as syndrome negative controls who do not have a positive centralized research test for SARS-CoV-2 will be classified as syndrome negative controls for the analysis ("analytical syndrome negative controls"). Patients enrolled as syndrome-negative controls who have a positive centralized research RT-PCR positive for SARS-CoV-2 will be excluded from the analysis. Classification criteria are detailed in Table 3.

**Table 3.** Classification of cases and control for analysis.

| **Status of patient at enrollment** | **Clinical SARS-CoV-2 test results** | **Research SARS-CoV-2 RT-PCR results** | **Classification for Analysis** |
|---|---|---|---|
| Enrollment case | ≥1 positive | Positive | Analytical case |
| Enrollment case | ≥1 positive | Negative | Analytical case |
| Enrollment case | ≥1 positive | Not done / inconclusive | Analytical case |
| Enrollment test-negative control | All negative | Positive | Analytical case |
| Enrollment test-negative control | All negative | Negative | Analytical test negative control |
| Enrollment test-negative control | All negative | Not done / inconclusive | Analytical test negative control |
| Syndrome negative control | Not done / inconclusive | Positive | Excluded |
| Syndrome negative control | Not done / inconclusive | Negative | Analytical syndrome negative control |
| Syndrome negative control | Not done / inconclusive | Not done / inconclusive | Excluded |
| Syndrome negative control | Negative | Positive | Excluded |
| Syndrome negative control | Negative | Negative | Analytical syndrome negative control |
| Syndrome negative control | Negative | Not done / inconclusive | Analytical syndrome negative control |

#### *11.1.ii Classification of vaccination status*

Enrolled patients who completed the full recommended vaccine regimen for the type of vaccine received at least 14 days prior to illness onset (or prior to hospital admission for syndrome negative controls) will be classified as fully vaccinated for the primary analysis. For example, for vaccines with 2 doses, a patient would be considered fully vaccinated 14 days after receiving the second dose. Patients who received some but not the full vaccine regimen at least 14 days prior to illness onset (or prior to hospital admission for syndrome negative controls) will be considered partially vaccinated.

#### *11.1.iii Primary and secondary models*

In the primary model, the adjusted odds ratio (aOR) of full SARS-CoV-2 vaccination will be calculated for cases compared to test-negative controls using multivariable logistic regression while adjusting for potential confounders. SARS-CoV-2 vaccine effectiveness for the prevention of COVID-19 hospitalizations will be calculated as (1 – aOR) x 100. Propensity score analysis may be used in a secondary vaccine effectiveness analysis.

In a secondary model using the syndrome negative control group, the adjusted odds ratio of full SARS-CoV-2 vaccination will be calculated for cases compared to syndrome-negative controls using multivariable logistic regression while adjusting for potential confounders. SARS-CoV-2 vaccine effectiveness will be calculated as (1-aOR) x 100.

#### *11.1.iv Additional vaccine effectiveness analyses*

In a series of additional analysis, we will estimate vaccine effectiveness for the prevention of severe manifestations of SARS-CoV-2 infection, by specific vaccine type, by SARS-CoV-2 variant, and by baseline patient characteristics (Table 4).

| **Table 4.** SARS-CoV-2 vaccine effectiveness analyses | |
|---|---|
| **Type of Analysis** | **Categories (levels)** |
| By fully vs partially vaccinated | Fully vaccinated (primary model)<br>Fully or partially vaccinated<br>Partially vaccinated |
| By different control groups | Test-negative control (primary model)<br>Syndrome-negative control |
| By severity of COVID-19 | All hospitalizations (primary model)<br>Hospitalization outside ICU<br>Hospitalization in ICU<br>Hospitalization with acute organ failure<br>Hospitalization with death<br>Severity defined by other parameters |
| By SARS-CoV-2 vaccine type | All vaccine types (primary model)<br>Pfizer vaccine<br>Moderna vaccine<br>TBD #1<br>TBD #2 |
| By SARS-CoV-2 variant | All SARS-CoV-2 (primary model)<br>Variant #1<br>Variant #2<br>Variant #3 |
| By patient age group | All adults (primary model)<br>Age 18-50 years<br>Age 50-65 years<br>Age 65-75 years<br>Age ≥75 years |
| By patient race/ethnicity | All race/ethnicity (primary model)<br>Non-Hispanic White<br>Non-Hispanic Black<br>Hispanic<br>Asian<br>Other |
| By patient comorbidity burden | All patients (primary model)<br>No chronic medical illnesses<br>1 chronic medical illness<br>2 chronic medical illnesses<br>≥3 chronic medical illnesses<br>Immunosuppression<br>Diabetes mellitus<br>Obesity<br>Other categories of chronic medical conditions |

#### 11.2 Duration of Protection from SARS-CoV-2 Vaccination

To assess the duration of protection from SARS-CoV-2 vaccines for the prevention of COVID-19 hospitalizations, we will collect the date of vaccination for participants, the date of onset for COVID-19 illness, the date of first positive test for SARS-CoV-2, and date of hospitalization. Using these data, we will describe the duration of time between vaccination and onset of COVID-19 for participants with "vaccine failure," defined as hospitalization for COVID-19 despite vaccination. Then, we will develop models to estimate duration of protection using vaccination status and duration of being vaccinated prior to illness onset while accounting for patient factors (age, comorbidities, long-term care facility resident) and pandemic factors (prevalence of cases in the community, uptake of vaccination in the community).

#### 11.3 Clinical Validity of SARS-CoV-2 RT-PCR Results

We will assess the agreement between SARS-CoV-2 RT-PCR tests obtained clinically at the time of hospital admission and other methods of diagnosing SARS-CoV-2 infection in approximately 250 enrolled patients. These will be the same patients selected for convalescent biospecimen sample collection, which will include both cases and patients enrolled as test-negative controls (patients hospitalized with acute respiratory illness and negative clinical SARS-CoV-2 testing). All these patients will have clinically obtained RT-PCR for SARS-CoV-2. We will compare the admission SARS-CoV-2 clinical RT-PCR result against three other variables: (1) SARS-CoV-2 RT-PCR results from a separate nasal sample collected by research personnel at the time of IVY enrollment and run at the Vanderbilt central IVY laboratory using CDC primers and probes; (2) SARS-CoV-2 antibody testing completed on acute and convalescent serum; (3) a clinical diagnosis (COVID-19 yes/no) based on chart review blinded to SARS-CoV-2 RT-PCR results from two clinicians with experience caring for COVID-19 patients.

This analyses will facilitate the comparison of a clinically obtained SARS-CoV-2 RT PCR results (which is what is being used to guide clinical care and a key component for vaccine effectiveness studies that rely on clinical testing) with three "criterion standards": (1) another RT-PCR conducted using alternative methods (inter-test reliability); an alternative laboratory method of diagnosing acute SARS CoV-2 infection (differential antibody detection in acute and convalescent serum); and (3) a clinical method of diagnosing acute SARS-CoV-2 infection.

### 12 Human Subjects

#### 12.1 Public Health Surveillance

This project will be conducted as non-research public health surveillance in accordance with the Code of Federal Regulations (CRF) and guidance from the Office for Human Research Protections (OHRP).

During the conduct of this project, we will collect information and biospecimens requested by CDC to monitor the ongoing COVID-19 pandemic. This project is supported and requested by CDC, which serves as the public health authority for this project. Activities will be limited to those necessary for CDC to monitor COVID-19 for the purposes of public health.

The criteria for non-research public health surveillance activities are outlined in 45 CFR 46.102(*l*)(2):

> "Public health surveillance activities, including the collection and testing of information or biospecimens, conducted, supported, requested, ordered, required, or authorized by a public health authority. Such activities are limited to those necessary to allow a public health authority to identify, monitor, assess, or investigate potential public health signals, onsets of disease outbreaks, or conditions of public health importance (including trends, signals, risk factors, patterns in diseases, or increases in injuries from using consumer products). Such activities include those associated with providing timely situational awareness and priority setting during the course of an event or crisis that threatens public health (including natural or man-made disasters)."

OHRP provides the following three criteria for an activity to be considered public health surveillance[6]:

- The activity must be a public health surveillance activity (45 CFR 46.102(*l*)(2));
- The activity must be conducted, supported, requested, ordered, required, or authorized by a public health authority (45 CFR 46.102(k) and 46.102(*l*)(2)); and
- The activity must be limited to that necessary to allow a public health authority to identify, monitor, assess, or investigate potential public health signals, onsets of disease outbreaks, or conditions of public health importance (including trends, signals, risk factors, patterns in diseases, or increases in injuries from using consumer products) (45 CFR 46.102(*l*)(2)).

#### 12.2 Collection and Storage of Health Information

During this surveillance activity, personnel will obtain health information from patient interview, surrogate interview, and medical records review. This will include personal information, including date of birth and date of hospitalization. Health information will be entered by personnel at each site into a REDCap electronic database maintained at Vanderbilt University Medical Center.

REDCap is a web-based data entry program accessible via an Internet connection. REDCap contains customizable data entry forms for data collection as well as an audit trail for tracking all activity within the system. REDCap is a secure web application for building and managing online surveys and databases. While REDCap can be used to collect virtually any type of data (including 21 CFR Part 11, FISMA, and HIPAA-compliant environments), it is specifically geared to support online or offline data capture for research studies and operations. REDCap employs several layers of security, including authentication of end-users, automatic user logout after 30 minutes of inactivity, a time-stamped audit trail, encrypted web-based information transmission, and firewall protection of uploaded documents. Additional information on REDCap is available at: www.project-redcap.org.

The primary risk of collection and storage of health information is inadvertent disclosure of this information. Inadvertent disclosure of health information will be minimized by using only professionals trained in good clinical practice and use of the REDCap system for data collection and storage. All personnel will have received appropriate training and certification in protection of subjects at their institutions.

#### 12.3 Collection and Storage of Biological Specimens

Enrolled patients may undergo collection of biological samples, including respiratory samples (such as, nasal swabs) and blood samples (such as, serum and plasma). Samples will be labeled with study numbers and linked to epidemiologic data collected from the patients via the study number. Samples will be shipped to Vanderbilt University Medical Center or CDC, where they will undergo analysis and long-term storage.

INFLUENZA AND OTHER VIRUSES IN THE ACUTELY ILL

Statistical Analysis Plan

**Adult Inpatient SARS-CoV-2 Vaccine Effectiveness Surveillance**

| | |
|---|---|
| Full Title | Adult Inpatient SARS-CoV-2 Vaccine Effectiveness Surveillance |
| IVY protocol number | IVY-3 |
| Short title | IVY-3 SARS-CoV-2 VE |
| Statistical Analysis Plan No. | 1 |
| Date | May 14, 2021 |
| Funder | Centers for Disease Control and Prevention (CDC) |
| Network | Influenza and Other Viruses in the Acutely Ill (IVY) |
| Coordinating Center | Vanderbilt University Medical Center |
| Analytical Lead | Mark Tenforde, MD, PhD<br>Centers for Disease Control and Prevention (CDC) |
| Lead Scientists | Wesley H. Self, MD, MPH<br>Vanderbilt University Medical Center<br>Manish M. Patel, MD<br>Centers for Disease Control and Prevention (CDC)<br>Jennifer R. Verani, MD, MPH<br>Centers for Disease Control and Prevention (CDC) |

**Statistical Analysis Plan: Table of Contents**

**I. Study Design**

Case-control investigations will be conducted to estimate COVID-19 VE against hospitalized illness by comparing odds of prior COVID-19 vaccination among adults hospitalized with illness consistent with COVID-19 who test positive for SARS-CoV-2 (cases) and hospitalized adults who test negative for SARS-CoV-2 (controls). Vaccination coverage in the control groups is meant to represent background coverage in similar adults from the study sites to compare against coverage in laboratory-confirmed cases. Two hospital controls groups will be included for these analyses, "test-negative" and "syndrome-negative" controls. For primary VE analyses, adults hospitalized with an illness consistent with COVID-19 who test negative for SARS-CoV-2 will be used as "test-negative" controls.

Delayed presentation to medical care and reduced sensitivity of SARS-CoV-2 diagnostic tests over time following infection may result in misclassification of some cases as controls (i.e., false-negatives). Therefore, a secondary "syndrome-negative" control group will also be used to assess VE. This control group will include hospitalized adults who do not have signs or symptoms consistent with COVID-19 and test negative for SARS-CoV-2 and are thus unlikely to be hospitalized for COVID-19-related illness.

**II. Eligibility criteria**

Eligibility criteria for test-positive cases (Cohort 1), test-negative controls (Cohort 2), and syndrome-negative controls (Cohort 3) are included below.

***Cohort 1: SARS-CoV-2 Cases***

Summary for Cohort 1: Adult admitted to the hospital for acute COVID-19 who has tested positive for SARS-CoV-2.

<u>Inclusion for Cohort 1 (cases):</u>

1. Age ≥18 years old.
2. Hospital admission or in an emergency department awaiting hospital admission.
3. Symptoms and/or signs believed to be due to COVID-19, including at least 1 of the following: fever; cough; shortness of breath; loss of taste; loss of smell; use of respiratory support (high flow oxygen by nasal cannula, non-invasive ventilation or invasive ventilation) for the acute illness; new pulmonary findings on chest imaging consistent with pneumonia.
4. Clinically obtained test that is positive for acute SARS-CoV-2 infection. The positive test may be obtained before or after hospital arrival. Examples of acute SARS-CoV-2 tests include RT-PCR tests, nucleic acid amplification tests (NAAT), and antigen tests. Serology testing may not be used for eligibility.

<u>Exclusion for Cohort 1 (cases):</u>

1. Previous enrollment as a case in this study.
2. The first positive test for acute SARS-CoV-2 infection is known to have occurred more than 10 days after onset of COVID-19 symptoms/signs listed in inclusion criterion #3. Patients with unknown onset date for COVID-19 symptoms/signs may be enrolled.
3. Hospital presentation for the COVID-19 admission is known to have occurred more than 14 days after onset of COVID-19 symptoms/signs listed in inclusion criterion #3. Patients with unknown onset date for COVID-19 symptoms/signs may be enrolled. Patients transferred from other

hospitals may be enrolled; the time of hospital presentation is the time of presentation to the first hospital.

***Cohort 2: SARS-CoV-2 Test Negative Controls***

Summary for Cohort 2: Adult admitted to the hospital for an acute illness with symptom overlap with COVID-19 who has tested negative for SARS-CoV-2.

<u>Inclusion for Cohort 2 (test-negative controls):</u>

1. Age ≥18 years old.
2. Hospital admission or in an emergency department awaiting hospital admission.
3. Symptoms and/or signs that overlap with COVID-19, including at least 1 of the following: fever; cough; shortness of breath; loss of taste; loss of smell; use of respiratory support (high flow oxygen by nasal cannula, non-invasive ventilation or invasive ventilation) for the acute illness; new pulmonary findings on chest imaging consistent with pneumonia.
4. Clinically obtained test that is negative for acute SARS-CoV-2. The negative test may be obtained before or after hospital arrival. Examples of acute SARS-CoV-2 tests include RT-PCR tests, NAAT, and antigen tests. Serology testing may not be used for eligibility.

<u>Exclusion for Cohort 2 (test-negative controls):</u>

1. Previous enrollment as a control in this study.
2. The first negative test for acute SARS-CoV-2 infection is known to have occurred more than 10 days after onset of symptoms/signs listed in inclusion criterion #3. Patients with unknown onset date for COVID-19 symptoms/signs may be enrolled.
3. Hospital presentation for the admission is known to have occurred more than 14 days after onset of symptoms/signs listed in inclusion criterion #3. Patients with unknown onset date for symptoms/signs may be enrolled. Patients transferred from other hospitals may be enrolled; the time of hospital presentation is the time of presentation to the first hospital.
4. Any positive test for acute SARS-CoV-2 infection in the 14 days prior to hospital presentation or between hospital presentation and enrollment (patients with a positive acute SARS-CoV-2 test should be screened for potential enrollment as a case).

***Cohort 3: SARS-CoV-2 Syndrome Negative Controls***

Summary for Cohort 3: Adult admitted to the hospital for a reason other than an acute respiratory illness and who does not have a clinical suspicion for COVID-19.

<u>Inclusion for Cohort 3 (syndrome-negative controls):</u>

1. Age ≥18 years old.
2. Hospital admission or in an emergency department awaiting admission.
3. Clinical impression that COVID-19 is not the reason for admission.

4. None of the following signs or symptoms that overlap with COVID-19 in the past 14 days: fever; cough; shortness of breath; loss of taste; loss of smell; use of respiratory support (high flow oxygen by nasal cannula, non-invasive ventilation or invasive ventilation) for the acute illness; new pulmonary findings on chest imaging consistent with pneumonia.

Exclusion for Cohort 3 (syndrome-negative controls):

1. Previous enrollment as a control in this study.
2. Any positive test for acute SARS-CoV-2 infection in the 14 days prior to hospital presentation or between hospital presentation and enrollment (patients with a positive acute SARS-CoV-2 test should be screened for potential enrollment as a case).

### III. Key objectives

1. Estimate vaccine effectiveness for the COVID-19 vaccines in use in the United States to prevent COVID-19 hospitalization among adults.
2. Estimate COVID-19 vaccine effectiveness for full vaccination (completion of 1-dose or 2-dose series) or partial vaccination (completion of only 1 dose in a 2-dose series).
3. Estimate COVID-19 vaccine effectiveness for the different SARS-CoV-2 vaccines in use in the United States for prevention of COVID-19 hospitalizations.
4. Estimate COVID-19 vaccine effectiveness against circulating SARS-CoV-2 variants for prevention of COVID-19 hospitalizations.
5. Estimate COVID-19 vaccine effectiveness for prevention of COVID-19 hospitalizations within subgroups of the population defined by age, race/ethnicity, and comorbidities (e.g., immunocompromising conditions).
6. Assess the duration of protection for COVID-19 vaccination.
7. Additional objectives may be added over time.

**IV. Sample size calculations**

Table 1 provides the estimated number of cases and test-negative controls required for a crude VE analysis using a range of assumptions for VE, precision (95% confidence interval), and vaccine coverage among control patients.

**Table 1**. Sample size calculators for VE analysis

| VE estimate | Confidence interval | Vaccine coverage in controls | Number of cases | Number of test-negative controls |
|---|---|---|---|---|
| 90% | 67.0–97.0% | 70% | 30 | 30 |
| 90% | 67.0–97.0% | 50% | 43 | 43 |
| 90% | 67.0–97.0% | 30% | 81 | 81 |
| 80% | 60.0–90.0% | 70% | 75 | 75 |
| 80% | 60.0–90.0% | 50% | 90 | 90 |
| 80% | 60.0–90.0% | 30% | 148 | 148 |
| 70% | 51.5–81.5% | 70% | 147 | 147 |
| 70% | 51.5–81.5% | 50% | 160 | 160 |
| 70% | 51.5–81.5% | 30% | 243 | 243 |

**V. Key study definitions**

Key study definitions for the VE analysis and clinical outcomes for specified severity outcomes are shown in Table 2A and 2B below:

**Table 2A. Study definitions.**

| **Category / Group** | **Description** |
|---|---|
| **Vaccination status** | |
| Full vaccination | Study patient who received all doses of an authorized COVID-19 vaccine ≥14 days prior to illness onset (or date of hospitalization in syndrome-negative controls). |
| Partial vaccination | Study patient who received one of two doses of a two-dose COVID-19 vaccine ≥14 days prior to illness onset (or date of hospitalization in syndrome-negative control patients). These patients either have not received a second dose of vaccine or received the second dose 0-13 days prior to illness onset. Secondary analyses may also consider VE within different timeframes from date of vaccination. |
| Full or partial vaccination (one or more doses) | Study patient who has received either full or partial vaccination with a COVID-19 vaccine. |
| Unvaccinated | Study patient who did not receive any doses of an authorized COVID-19 vaccine ≥14 days prior to illness onset or who was vaccinated after illness onset. A patient who received vaccination 0-13 days before illness onset will be excluded from the primary VE analyses but will be considered in secondary analyses. This vaccination group may serve as an indicator of bias in primary VE analyses (selection bias or confounding not accounted for in VE models), as vaccine effectiveness is not expected in the period shortly following the first dose of a COVID-19 vaccine. |
| **Alternative times from vaccination to illness onset** | If feasible we may perform additional sub-analyses of VE using other discrete time periods between vaccination and illness onset, such as (for a 2-dose vaccine series): |

| | |
|---|---|
| | 0-14 days from first dose of vaccine (partial vaccination)<br>15-28 days from first dose of vaccine (partial vaccination)<br>29-42 days from first dose of vaccine (partial vaccination)<br>>42 days from first dose of vaccine (partial vaccination)<br>0-7 days from second dose of vaccine<br>8-14 days from second dose of vaccine<br>>14 days from second dose of vaccine<br><br>Future analyses will evaluate potential waning vaccine effectiveness over time in future analyses, e.g. for each 30-day interval from date of vaccination. |
| **Severity** | |
| Severe case | Several definitions for severity will be used including but not limited to case patients who are: hospitalized in ICU, hospitalized with acute organ failure, hospitalized with vasopressor-dependent shock, hospitalized with death, hospitalized with non-invasive or invasive mechanical ventilation, or severity defined by other parameters (e.g., Food and Drug Association criteria). |
| Non-severe case | Case patient who do not meet criteria for the severity case definition. |
| **Genetic strain** | |
| SARS-CoV-2 strain | Available specimens with a cycle threshold (Ct) value ≤30 will undergo genetic sequencing with lineage determination. VE by group (B.1.1.7, B.1.351, or P.1 viruses, or other variants) will be assessed if sufficient sample size. For specimens with higher Ct values, PCR analysis using targeted primers to detect SARS-CoV-2 strains will be conducted. |

**Table 2B. Clinical outcome definitions**

| Clinical Outcome | Description |
|---|---|
| **Window of outcome assessment** | All outcomes are assessed between initial hospital presentation and the earlier of hospital discharge or the end of hospital day #28 (calendar day). Once a patient is discharged from the acute care hospital participating, outcome assessment ends. Clinical events, such as vasopressor use and mechanical ventilation that occur during the index hospitalization but before enrollment (defined as the time of upper respiratory sample collection) are included as study outcomes. |
| **ICU admission** | Admission to or boarding for an ICU bed at any time between initial hospital presentation and the earlier of discharge or hospital day #28. |
| **Vasopressor support** | Use of midodrine or a vasopressor by continuous intravenous/intraosseous infusion for at least 1 hour to increase or maintain blood pressure between initial hospital presentation and the earlier of hospital discharge or the end of hospital day #28. Vasopressors may include angiotensin II, isoproterenol, epinephrine, midodrine, norepinephrine, dopamine, phenylephrine, vasopressin. Continuation of chronic outpatient midodrine dosing does not fulfill criteria for vasopressor-dependent shock. |
| **Invasive mechanical ventilation** | Receipt of positive pressure ventilation through an endotracheal tube or tracheostomy tube for at least 1 hour between initial hospital presentation and the earlier of hospital discharge or the end of hospital day #28. Patients on chronic invasive mechanical ventilation prior to the current illness are not eligible for the invasive mechanical ventilation outcome. |

| | |
|---|---|
| **Non-invasive ventilation** | Receipt of positive pressure ventilation through nasal prongs or a facemask for at least 1 hour between initial hospital presentation and the earlier of hospital discharge or the end of hospital day #28. High flow nasal cannula, CPAP, and BiPAP for respirator support, other than for treatment of chronic sleep apnea, fulfill the criteria for the non-invasive mechanical ventilation outcome. Patients chronically on invasive or non-invasive mechanical ventilation prior to the current illness are not eligible for the non-invasive mechanical ventilation outcome. |
| **In-hospital 28-day mortality** | Death after hospital presentation and before the earlier of hospital discharge or the end of hospital day #28. |

### VI. Vaccine Effectiveness Analyses

#### Vaccination status definition (exposure)

COVID-19 vaccine verification will occur using a systematic process. Patients or their proxies are interviewed about receipt of one or more doses of a COVID-19 vaccine. If no COIVD-19 vaccine is self-reported or vaccine history cannot be completed, local hospital EMR and vaccine registry searches will be completed (first search shortly after enrollment and second search approximately 28 days after enrollment). If receipt of at least one COVID-19 vaccine is self-reported, vaccine verification will continue until all options for verification have been exhausted and the vaccine dose(s) cannot be verified. When all reported vaccine doses have been verified, the vaccine verification process will stop. Sources of documentation include CDC vaccine card, local hospital EMR, state vaccine registry (including search shortly after enrollment and approximately 28 days after enrollment), and vaccine records requested from other sources including clinics or pharmacies (if the patient reported receiving a vaccine at one of these sources and receipt cannot be verified using the above sources).

Patients vaccinated after illness onset (or hospital admission in the syndrome-negative control group) will be classified as unvaccinated, and patients who received a first dose of a licensed SARS-CoV-2 vaccine 0-13 days before illness onset will be excluded from the primary analysis given unlikely vaccine-associated protection during this interval shortly following vaccination.

Definition 1: Documented vaccination
Requires documented evidence of vaccination, operationalized as a non-missing date of vaccination obtained from a vaccination record card, electronic medical record (EMR), local vaccine registry, or other documented source (including other clinics or pharmacies).

Definition 2: Self-reported vaccination with date
Requires patient/proxy to answer affirmatively to yes/no receipt of COVID-19 vaccine AND be able to provide either an exact or approximate date of vaccination, in order to establish that the vaccine was received at least two weeks prior to onset. If patient cannot answer that question, they are classified as having missing date of vaccination and self-reported vaccination is also classified as missing/unknown.

Definition 3: Self-reported vaccination with date and location ("plausible self-report")
Same as definition 2 except that patient/proxy must also verbally provide a location of vaccination. Usually, any reasonable location is accepted.

Definition 4: Self-report with date OR documented
Classified as vaccinated if patient meets either definition 1 or definition 2

Definition 5: Plausible self-report OR documented
Classified as vaccinated if patient meets either definition 1 or definition 3

**SARS-CoV-2 case status definitions (outcome)**

Patients will be enrolled into Cohort 1, Cohort 2, or Cohort 3. New or stored upper respiratory specimens or salivary specimens will be routinely collected from all enrolled patients for centralized research RT-PCR testing. Patients enrolled in Cohort 2 who had negative clinical SARS-CoV-2 testing will be reclassified as an “analytical case” if RT-PCR testing is positive. Patients enrolled in Cohort 3 will be excluded from the analysis if research RT-PCR testing is positive for SARS-CoV-2.

Classification of cases and controls for the analysis is shown below in Table 3:

**Table 3.** Classification of cases and control for analysis.

| **Status of patient at enrollment** | **Clinical SARS-CoV-2 test results** | **Research SARS-CoV-2 RT-PCR results** | **Classification for Analysis** |
|---|---|---|---|
| Enrollment case | ≥1 positive | Positive | Analytical case |
| Enrollment case | ≥1 positive | Negative | Analytical case |
| Enrollment case | ≥1 positive | Not done / inconclusive | Analytical case |
| Enrollment test-negative control | All negative | Positive | Analytical case |
| Enrollment test-negative control | All negative | Negative | Analytical test negative control |
| Enrollment test-negative control | All negative | Not done / inconclusive | Analytical test negative control |
| Syndrome negative control | Not done / inconclusive | Positive | Excluded |
| Syndrome negative control | Not done / inconclusive | Negative | Analytical syndrome negative control |
| Syndrome negative control | Not done / inconclusive | Not done / inconclusive | Excluded |
| Syndrome negative control | All negative | Positive | Excluded |
| Syndrome negative control | All negative | Negative | Analytical syndrome negative control |
| Syndrome negative control | All negative | Not done / inconclusive | Analytical syndrome negative control |

**Exclusion criteria for analysis:**

- Missing/unknown case status
- Missing/unknown vaccination status
- First dose of vaccine 0 to 13 days before illness onset (excluded from primary analysis)
- Illness onset date after admission (to exclude potential nosocomial infections or other reasons for admission)
- Illness onset >10 days before date of first SARS-CoV-2 test

- Illness onset >14 days before date of hospitalization
- Received a COVID-19 vaccine not authorized for use in the United States

**VII. Model building strategy for final VE analysis**

*Patient description:*

A model building strategy will be applied for VE assessment using test-negative or syndrome-negative control groups. Characteristics of SARS-CoV-2-positive and SARS-CoV-2-negative patients (or syndrome-negative controls) and vaccinated and unvaccinated patients will be described by counts and percentages or medians and interquartile ranges and compared using Pearson $X^2$ test or Fisher exact test for categorical variables and Wilcoxon rank-sum test or *t* test for continuous variables.

*Adjusted COVID-19 VE:*

Crude (unadjusted) VE will first be calculated. Adjusted VE will then be calculated by comparing the odds of COVID-19 vaccine receipt among cases and controls using unconditional multivariable logistic regression with SARS-CoV-2 positivity as the outcome and vaccination status as the exposure. VE is calculated as (1 – *adjusted odds ratio*) × 100%.

Several variables will be included *a priori* in VE regression models. These include enrolment location (specific hospital or geographic region), age, sex, race/ethnicity, and calendar time. These variables are included because they are established or suspected confounders.

Additional variables will be considered as model covariates that may confound the relationship of interest if associated with both vaccination status and risk of hospitalized COVID-19 illness.

We will examine the association of these other potential confounders. Variables with the strongest association with the exposure and the outcome that change the OR by >5-10% will be selected for inclusion in the model. If several of these potential confounders performed similarly, we will use the most parsimonious set of variables. We will examine a range of health status indicators as potential confounders, including measures of baseline health status (e.g., hospitalizations in the past year). As factors related to social vulnerability and community exposures/behaviors are also potential confounders, we will also consider measures of SES and social vulnerability (e.g., CDC Social Vulnerability Index) and reported behaviors/exposures (e.g. long-term care facility resident, healthcare worker, influenza vaccination during the current season).

We will estimate VE separately for full vaccination, partial vaccination, and receipt of one or more doses (either full or partial vaccination), as well as for subgroups of interest or using alternative times since vaccination. VE will also be estimated based on documented vaccination alone (Definition 1 above) as well as either documented vaccination or plausible self-report including location and date(s) [Definition 5 above]. Patients self-reporting COVID-19 vaccination without details about location and date(s) will be excluded from the analysis given low confidence in vaccination status. Similar approaches to model building will also be used in estimating VE using the syndrome-negative control group.

*Effect modification:*

Potential effect modifiers of vaccine effectiveness, such as presence of immunocompromising conditions or history of prior SARS-CoV-2 infection, may also be considered. Likelihood ratio tests will be used to compare P-values of the interaction term (a P-value <~0.15 is suggestive of effect modification). VE estimates will be presented stratified by level of an effect modifier.

*Propensity score sensitivity analysis:*
Main VE analyses will be performed using unconditional logistic regression analysis, with confounders included as covariates to reduce bias. Propensity score analysis, a quasi-experimental observational method, will be considered as a sensitivity approach to determining VE should significant imbalance be observed between vaccinated and unvaccinated groups. Propensity scores represent the probability of exposure (i.e., SARS-CoV-2 vaccination) conditioned on observed baseline characteristics. Propensity scores function by making the exposure independent of confounders through balancing confounders between groups during the analysis phase.

**VIII. Other analytic considerations**

*Missing illness onset date:*

Some patients or proxies may not be able to provide the date of illness onset. For the main VE analysis, we will include these patients. We may perform sensitivity analyses restricting the VE analysis only to patients with a known date of illness onset, using a multiple imputation approach to impute illness onset date in reference to time of testing for those with missing values, or assigning illness onset date as the median days between illness onset and date of hospital admission in patients with complete data.

*Specification of race/ethnicity:*

We may be unable to obtain race and ethnicity data for all patients. This will be handled analytically, e.g., including an unknown race/ethnicity category if this group is large.

*Specification of age:*

Specification of age varies depending on subset being analyzed. Alternative specifications included continuous/linear (if data suggested no evidence of nonlinearity) or 3-or 4-part categorization if nonlinearity is present (alternatively, cubic splines can be used). In models stratified by age group, age (continuous) is typically included in the model to account for residual confounding by age within age category (sample size permitting).

*Specification of calendar time:*

Calendar time of symptom onset is specified in the VE model by classifying based on calendar week based on the date of illness onset or date of hospitalization. We will consider adjusting for calendar time in different intervals (e.g., weekly, biweekly).

*Discordant dates of illness onset or dates of vaccination:*

If patient reports a date of illness onset inconsistent with date of onset from the electronic medical record (EMR), we generally take the patient self-reported value. However, dates of COVID-19 vaccination through documented sources such as vaccine record cards or immunization registries are likely to be more accurate than self-reported dates and will therefore be used preferentially.

### IX. Changes between original and final analysis plan (June 2, 2021)

The original statistical analysis plan (SAP) was finalized on May 14, 2021, prior performing statistical modeling or viewing summary results. During analysis, between May 14 and June 2, 2021, the following updates were made to the analysis plan. These modifications were made with knowledge of summary data in the first analyzed cohort (patients enrolled March 11 – May 5, 2021).

1. In the original SAP, the test-negative control group is listed as a primary control group and syndrome-negative control group as a secondary control group. In the final analysis, we estimate SARS-CoV-2 vaccine effectiveness separately for the test-negative and syndrome-negative control groups. Given highly similar vaccine effectiveness estimates with these individual control groups, we also combined patients from these two control groups to improve estimate precision as well as for clarity in presenting estimates.
2. In the original SAP, we state that we will exclude patients who received a SARS-CoV-2 vaccine not authorized for use in the United States. In the final analysis, we also excluded patients who had received the Jannsen Johnson & Johnson SARS-CoV-2 vaccine due to very low sample size of patients who received this vaccine.
3. In the original SAP, we state that variables with the strongest association with the outcome that change the adjusted odds ratio by >5-10% will be selected for inclusion in final models. In the final analysis, we considered covariates that generated a >5% absolute change in adjusted odds ratio in either direction. We list several covariates that were considered in adjusted models in the SAP, none of which changed the adjusted odds ratios appreciably (>5% absolute change) when added to base model.
4. In the original SAP, we state that we will consider current-season influenza vaccination as a co-variable in models but did not consider this covariate in the final analysis because information about prior season influenza vaccination was collected by self-report only without available source verification for most patients.
5. In the original SAP, we state that we will estimate vaccine effectiveness for patients who received one or more doses of vaccine (either partially or fully vaccinated). In the final analysis, we present separate vaccine effectiveness estimates for partial and full vaccination but do not present an estimate for combined partial or full vaccination. We decided not to present vaccine effectiveness for a combined partially or fully vaccinated group because in the initial analysis we had adequate sample size to estimate effectiveness for both partial and full vaccination separately. Furthermore, the interpretation of vaccine effectiveness for partial or full vaccination is not straight-forward.
6. In the original SAP, we state that available specimens with a SARS-CoV-2 target RT-PCR cycle threshold (Ct) value ≤30 will undergo genetic sequencing with lineage determination. In the final analysis, we changed the Ct threshold to ≤32 because the sequencing laboratory was able to obtain sequencing results from samples with a Ct up to 32.
7. In the original SAP, we state that we will formally compare baseline characteristics of vaccinated and unvaccinated patients using Pearson $X^2$ test or Fisher exact test for categorical variables and Wilcoxon rank-sum test or *t* test for continuous variables. In the final analysis, we present medians and interquartile ranges or numbers and percentages without presenting a formal comparison between groups.
8. In the final analysis, we performed a third sensitivity analysis not stated in the SAP, in which the reference date to determine vaccination status for the syndrome-negative control group was changed from 14 days prior to admission to the median number of days between illness onset and

admission for test-negative controls plus 14 days to more closely align with the method of reference determination for cases. Overall vaccine effectiveness estimates were very similar to those in the primary model.

9. In the original SAP, primary objective 5 states that an objective is to estimate COVID-19 vaccine effectiveness within subgroups as defined by certain comorbidities. Due to limited sample size of patients with certain co-morbidities, we only performed analyses for underlying medical conditions with >20% prevalence in the full study population.
